# Novel Deep Learning Framework for Simultaneous Assessment of Left Ventricular Mass and Longitudinal Strain: Clinical Feasibility and Validation in Patients with Hypertrophic Cardiomyopathy

**DOI:** 10.1101/2025.01.17.25320694

**Authors:** Jiesuck Park, Yeonyee E. Yoon, Yeonggul Jang, Taekgeun Jung, Jaeik Jeon, Seung-Ah Lee, Hong-Mi Choi, In-Chang Hwang, Eun Ju Chun, Goo-Yeong Cho, Hyuk-Jae Chang

**Affiliations:** Cardiovascular Center and Division of Cardiology, Department of Internal Medicine, Seoul National University Bundang Hospital, Seongnam, Gyeonggi, Republic of Korea; Department of Internal Medicine, Seoul National University College of Medicine, Seoul, Republic of Korea; Ontact Health Inc., Seoul, Republic of Korea; CONNECT-AI Research Center, Yonsei University College of Medicine, Seoul, Republic of Korea; Department of Radiology, Cardiovascular Center, Seoul National University Bundang Hospital, Seongnam, Gyeonggi-do, Republic of Korea; Department of Radiology, Seoul National University College of Medicine, Seoul, Republic of Korea; Division of Cardiology, Severance Cardiovascular Hospital, Yonsei University College of Medicine, Yonsei University Health System, Seoul, Republic of Korea

**Keywords:** cardiac magnetic resonance imaging, echocardiography, global longitudinal strain, hypertrophic cardiomyopathy, left ventricle, mass

## Abstract

**Background:** This study aims to present the Segmentation-based Myocardial Advanced Refinement Tracking (SMART) system, a novel artificial intelligence (AI)-based framework for transthoracic echocardiography (TTE) that incorporates motion tracking and left ventricular (LV) myocardial segmentation for automated LV mass (LVM) and global longitudinal strain (LVGLS) assessment.

**Methods:** The SMART system demonstrates LV speckle tracking based on motion vector estimation, refined by structural information using endocardial and epicardial segmentation throughout the cardiac cycle. This approach enables automated measurement of LVM_SMART_ and LVGLS_SMART_. The feasibility of SMART is validated in 111 hypertrophic cardiomyopathy (HCM) patients (median age: 58 years, 69% male) who underwent TTE and cardiac magnetic resonance imaging (CMR).

**Results:** LVGLS_SMART_ showed a strong correlation with conventional manual LVGLS measurements (Pearson’s correlation coefficient [PCC] 0.851; mean difference 0 [-2–0]). When compared to CMR as the reference standard for LVM, the conventional dimension-based TTE method overestimated LVM (PCC 0.652; mean difference: 106 [90–123]), whereas LVM_SMART_ demonstrated excellent agreement with CMR (PCC 0.843; mean difference: 1 [-11–13]). For predicting extensive myocardial fibrosis, LVGLS_SMART_ and LVM_SMART_ exhibited performance comparable to conventional LVGLS and CMR (AUC: 0.72 and 0.66, respectively). Patients identified as high-risk for extensive fibrosis by LVGLS_SMART_ and LVM_SMART_ had significantly higher rates of adverse outcomes, including heart failure hospitalization, new-onset atrial fibrillation, and defibrillator implantation.

**Conclusions:** The SMART technique provides a comparable LVGLS evaluation and a more accurate LVM assessment than conventional TTE, with predictive values for myocardial fibrosis and adverse outcomes. These findings support its utility in HCM management.

## INTRODUCTION

Transthoracic echocardiography (TTE) is the primary diagnostic tool for hypertrophic cardiomyopathy (HCM) [1, 2]. Evaluation of HCM typically involves assessing left ventricular (LV) hypertrophy and function through visual estimation and quantitative measurements, such as LV wall dimensions and ejection fraction (EF) [3]. However, thickened LV wall in HCM reduces cavity size, leading to overestimation of EF, making LV global longitudinal strain (LVGLS) a more accurate measure of myocardial contractility. Furthermore, dimension-based LV mass (LVM) calculation fails to accurately reflect diverse hypertrophic patterns seen in HCM [4, 5]. Consequently, cardiac magnetic resonance (CMR) is often required for a more precise evaluation [1], as LVM has prognostic significance in HCM, with higher values associated with adverse outcomes such as arrhythmia and sudden cardiac death [6, 7].

Recent advances in artificial intelligence (AI) have enabled the automated segmentation of LV cavity and wall, providing LV volume, EF, and mass in TTE [8–10]. Automated LVGLS measurement is also now available and validated across various populations [11–14]. However, the unique geometry of HCM complicates the automated detection of LV endocardial and epicardial borders, as well as accurate motion tracking. Consequently, validation of AI-based automated measurements remains limited in HCM, particularly in accurately measuring LVM in cases with asymmetric hypertrophy.

Our research group previously developed and validated a comprehensive automated system for TTE analysis [15–20], featuring an algorithm that simultaneously segments LV wall and tracks myocardial motion, which was successfully validated in myocardial infarction patients [19]. Building on this foundation, we developed the Segmentation-based Myocardial Advanced Refinement Tracking (SMART) technique, a novel approach that advances motion tracking by integrating LV myocardial segmentation throughout the cardiac cycle. In this study, we aimed to present the SMART technique for the first time and evaluate its ability to successfully segment and track LV wall in HCM patients, enabling accurate assessment of LVM (relative to CMR) and reliable measurement of LVGLS.

## MATERIALS AND METHODS

### Study Population

The AI-based system utilized in this study was developed and validated with data from the Open AI Dataset Project (AI-Hub), which was initiated by the Ministry of Science and ICT, Korea [15–21]. For the development of the SMART technique, no additional developmental datasets were required beyond those used in the previously developed automatic segmentation and strain analysis [19]. We conducted in-house validation of the SMART technique across diverse study populations, including the in-house test set, an American population, and a paired dataset with both TTE and CMR. (**Supplemental Method 1**)

In this study, we performed an independent external validation of the SMART technique using a completely separate cohort of HCM patients that had not been involved in the development or previous validation of SMART. Specifically, we utilized the same HCM patient cohort as described in our prior publication [22], but applied a distinct analytic framework to evaluate the feasibility and performance of our novel AI-based technique - SMART - for the simultaneous quantification of LVM and LVGLS in patients with HCM. Among 306 consecutive patients with HCM who underwent CMR at Seoul National University Bundang Hospital between 2010 and 2019, 116 also underwent TTE within ± 6 months of CMR. The clinical diagnosis of HCM was defined by a maximal end-diastolic wall thickness >15 mm in any LV segment, excluding other potential causes of hypertrophy. We excluded four patients with inadequate image quality for strain analysis (three TTE, one CMR) and one patient in whom late gadolinium enhancement (LGE) quantification was not feasible, resulting in a final study population of 111 patients.

The study protocol was approved by the Institutional Review Board of Seoul National University Bundang Hospital (IRB No. B-2305-827-002), with a waiver of informed consent granted due to the retrospective study design. All clinical data were fully anonymized prior to analysis. The study was conducted in accordance with the principles outlined in the Declaration of Helsinki (2013).

### TTE Acquisition and Analysis

All echocardiographic examinations were performed by trained echocardiographers or cardiologists and interpreted by board-certified cardiologists with expertise in echocardiography following current guidelines. Standard ultrasound machines (Vivid 7, E9, and E95; GE Vingmed Ultrasound AS, Norway) were used with a 2.5-MHz probe at 50 to 80 Hz frame rates. Following standard protocols, 2-dimensional (2D), M-mode, and Doppler images were obtained.

LV maximal wall thickness (LVMWT_TTE_) was measured at the thickest segment of LV wall in end-diastole (ED). Dimension-based LVM (LVM_TTE_-_DB_) was calculated using the interventricular septum (IVS), left ventricular internal diameter (LVID), and posterior wall thickness (PWT) in ED according to the Devereux equation [4]:

0.8× 1.04 × [(IVS + LVID + PWT)^3^ − LVID^3^] + 0.6 g

LVGLS analysis was performed by an echocardiography specialist with over 20 years of experience, using EchoPAC PC BT20 (GE Medical Systems, Norway) (LVGLS_TTE-EchoPAC_). The initial measurements were reviewed in consensus by two board-certified cardiologists with subspecialty expertise in echocardiography (YE Yoon, >15 years of experience; SA Lee, > 10 years of experience). Manual strain analysis was conducted prior to the SMART processing and was performed entirely independently of the CMR analysis. All echocardiographic images were saved as DICOM files and analyzed on dedicated workstations. The final LVGLS was derived by averaging the LVGLS from the apical 3-chamber (A3C), 4-chamber (A4C), and 2-chamber (A2C) views.

### CMR Acquisition and Analysis

The detailed processes for CMR imaging were provided in **Supplemental Methods 2**. The CMR images were acquired under electrocardiographic gating and breath-hold conditions. Steady-state free-precession cine-CMR images were obtained in the horizontal long axis, vertical long axis, and LV outflow tract. A short-axis stack view of the whole LV was obtained for LV volume and mass analysis. LGE images were obtained 10 minutes after intravenous administration of 0.2 mmol/kg of gadodiamide using a phase-sensitive inversion-recovery turbo field echo sequence.

All acquired images were processed using CVI42 software (version 5.10; Circle Cardiovascular Imaging, Calgary, Canada) and analyzed by an independent radiologist blinded to clinical and echocardiographic data. Endocardial and epicardial borders were auto-traced, with manual correction for contours with apparent deviation. LVM_CMR_ were estimated from short-axis cine-CMR. LVGLS analysis was conducted via CMR tissue-tracking (LVGLS_CMR_), which uses a mid-surface curvilinear coordinate system to track myocardial deformation and follows the motion of software-generated myocardial nodes on cine sequences [23]. Using long-axis cine-images, the whole myocardial LVGLS was calculated throughout the cardiac cycle, with the peak value designated as LVGLS_CMR_. LGE mass was quantified using the full-width at half-maximum method. The total LGE mass was obtained by summing the LGE from all sections, and the relative extent of LGE was expressed as a percentage of total LVM. Extensive LGE was defined as involving >15% of LVM, a threshold associated with an increased risk of sudden cardiac death (SCD) [1, 22, 24].

### Automatic TTE Analysis Using SMART Technique

Our AI-based system (Sonix Health Workstation, version 2.0; Ontact Health Inc., Korea) performs fully automated tasks, including view classification, segmentation, and echocardiographic parameter extraction [13, 18–20]. However, as TTE views for GLS measurement were pre-selected for comparison with conventional manual measurement of LVGLS, automatic view classification results were not used for validating the performance of the SMART technique in this study. Nevertheless, for completeness, automatic view classification performance was additionally assessed on the current dataset and demonstrated high accuracy (overall classification accuracy of 99.6%), as detailed in **Supplemental Method 3**.

The SMART technique is a fully automated deep learning (DL)-based method that integrates myocardial segmentation with speckle tracking to improve the accuracy of LV quantification from 2D TTE. A detailed description of the SMART architecture and computational framework is provided in **Supplemental Method 4**. This technique enables simultaneous measurement of key LV parameters - including LV end-diastolic and end-systolic volumes (LVEDV, LVESV), LVM, and LVGLS – without any operator input or manual correction. The process begins with motion vector estimation, followed by refinement using segmentation-based information (**Figure 1**). This involves a two-step raster-scan refinement of the region of interest (ROI): first, the mid-myocardial line is aligned to the myocardial structure; second, the endocardial and epicardial borders are corrected. This allows reliable speckle tracking and geometry extraction across the cardiac cycle.

**Figure 1.**
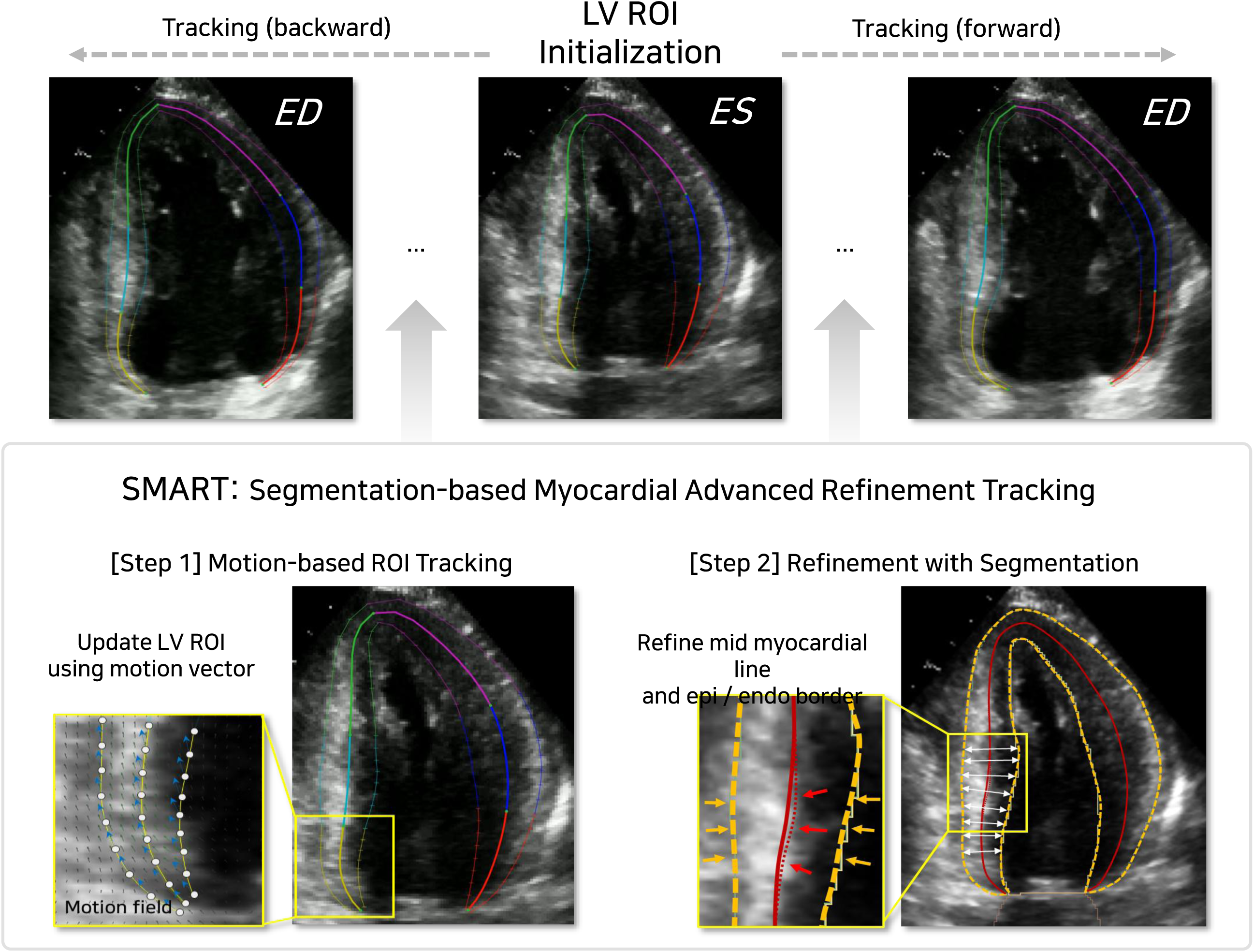
The SMART framework. The workflow highlights the dynamic update of LV ROI using a bi-directional dense motion field and a segmentation-guided refinement process to achieve myocardial structure-aware tracking for enhanced functional assessment. The process begins with segmentation of the LV myocardium across all frames, constructing an LV area curve for cardiac cycle analysis, including the identification of key phases such as ED and ES. Motion vectors are estimated and iteratively refined through a two-step process: first, by aligning the mid-myocardial line with the myocardial structure, and second, by precisely delineating the epicardial and endocardial borders. Abbreviations: ED, end-diastole; ES, end-systole; LV, left ventricle; ROI, region of interest; SMART, Segmentation-based Myocardial Advanced Refinement Tracking.

For each frame in the A4C, A2C, and A3C views, endocardial and epicardial borders are extracted, and geometric parameters such as LV radius and myocardial thickness at the basal, mid, and apical levels are calculated. These are then mapped into a tri-planar configuration (A4C-A2C-A3C) to reconstruct LV geometry (**Supplemental Method 4**). LV volume is derived from the endocardial border, and LVM (LVM_SMART_) is calculated as the difference between epicardial and endocardial volumes, multiplied by a myocardial tissue density of 1.05. All SMART-derived values used in this study were generated without operator intervention. Identical outputs would be produced for a given echocardiographic image, ensuring full reproducibility.

SMART processing is computationally efficient and suitable for standard clinical environments. Based on execution using a typical central processing unit (CPU) system (Intel Core i7-12th Gen 12700K, Alder Lake), segmentation and motion estimation are completed in under 6 seconds, phase detection and tracking in under 3 seconds, and LV volume and mass calculations in under 0.1 seconds per patient. Thus, full analysis is typically achieved within a few seconds, without the need for GPU acceleration. These features facilitate seamless integration of SMART into routine echocardiographic workflows (**Supplemental Method 5**). In-house validation results of the SMART technique are detailed in **Supplemental Methods 6, 7**.

### Statistical analysis

Continuous variables are presented as median (interquartile range) and categorical variables are reported as numbers (proportions). The concordance between automatic LVGLS measurements derived from the SMART technique and conventional manual measurements was assessed using the Pearson Correlation Coefficient (PCC) and mean absolute error (MAE). The agreement was further evaluated through Bland-Altman analysis, reporting the mean difference and limits of agreement. Similarly, LVMWT_TTE_, LVM_TTE-DB_, and LVM_SMART_ were compared against LVM_CMR_, the ground truth. Discrimination performance for extensive LGE (>15% LVM) was evaluated across different LVGLS, LVMWT and LVM methods using the area under the receiver operating characteristic curves (AUC). The statistical significance of the performance difference between SMART and conventional echocardiographic parameters was assessed using the DeLong test. Optimal cutoffs for LVGLS_SMART_ and LVM_SMART_ were obtained from receiver operating characteristics (ROC) curves to predict extensive LGE, enabling patient stratification. Clinical outcomes, including all-cause mortality, heart failure admission, new-onset atrial fibrillation (AF), and new implantable cardioverter-defibrillator (ICD) insertion, were analyzed as a composite endpoint. Survival curves and Cox regression analysis, adjusted for age, sex, body mass index, systolic blood pressure, LVEF, and e’ velocity, were used to compare outcome risks and calculate hazard ratios (HR). Statistical analyses were performed using R software (version 4.3.2; R Development Core Team, Vienna, Austria), with two-sided p-values <0.05 considered statistically significant.

## RESULT

### Baseline Characteristics

**Table 1** summarizes the baseline characteristics of 111 HCM patients (median age: 58 years, male 69%). Among them, 41 (37%) had mixed or diffuse type HCM, 37 (33%) had septal type HCM, and 33 (30%) had apical type HCM. Additionally, 16 patients (14%) exhibited extensive LGE. While age, sex, and BMI did not significantly differ between patients with and without extensive LGE, those with extensive LGE had a higher prevalence of mixed or diffuse HCM phenotypes.

When stratified by extensive LGE, LVGLS was significantly lower in patients with extensive LGE across all measurement methods, indicating more impaired LV function. This reduction was observed with EchoPAC (LVGLS_TTE-EchoPAC_, 11% vs. 14%, p=0.002), the SMART technique (LVGLS_SMART_, 11% vs. 14%, p=0.005), and CMR (LVGLS_CMR_, 10% vs 11%, p<0.001). Conventional TTE-derived LVM measures, such as LVMWT_TTE_ and LVM_TTE_-_DB_, showed no significant differences between patients with and without extensive LGE. However, LVM_SMART_ and LVM_CMR_ were significantly higher in patients with extensive LGE (161 g vs. 133 g; p=0.037, and 188 g vs. 123 g; p=0.007, respectively).

### Performance of SMART Technique

No failures of SMART analysis occurred; all included cases were successfully processed without operator intervention. The differences between methods for LVGLS measurement are summarized in **Table 2**. The correlation plot demonstrates LVGLS_SMART_ is well correlated with conventionally measured LVGLS_TTE-EchoPAC_ with high concordance (mean difference, 0 [-1 to 0]; p for difference=0.312) (**Figure 2**). When compared to LVGLS_CMR_, both LVGLS_SMART_ and LVGLS_TTE-EchoPAC_ showed significant differences (p for difference <0.001); however, discrepancies were small (mean difference, LVGLS_SMART_; 2 [1 to 3], and LVGLS_TTE_EchoPAC_; 3 [2 to 3]).

**Figure 2.**
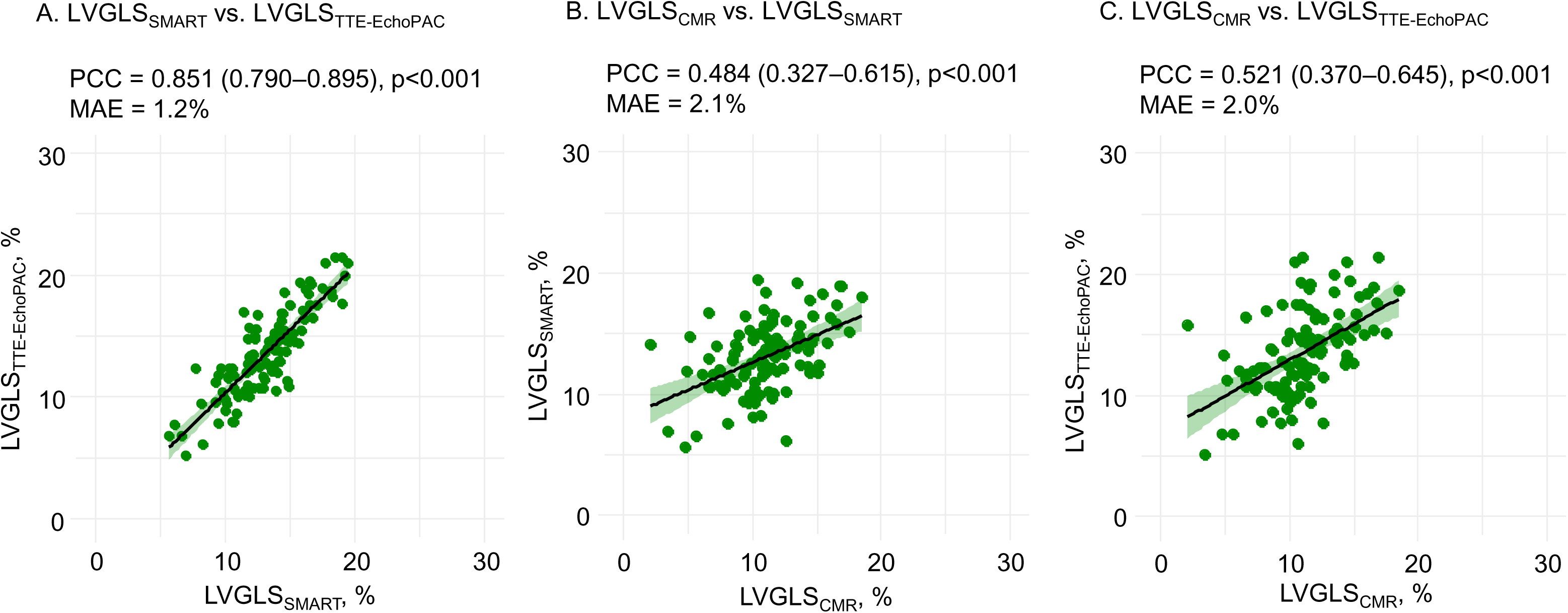
Concordance of LVGLS among SMART, manual measurement, and CMR The correlation plot demonstrated a significant association for LVGLS between SMART (LVGLS_SMART_) and manual measurements using EchoPAC (LVGLS_TTE-EchoPAC_) (A). When compared to CMR-based LVGLS (LVGLS_CMR_), both LVGLS_SMART_ (B) and LVGLS_TTE-EchoPAC_ (C) showed significant correlations. Abbreviations as in Figure 1: CMR, cardiovascular magnetic resonance imaging; LVGLS, left ventricular global longitudinal strain; MAE, mean absolute error; PCC, Pearson Correlation Coefficient; TTE, transthoracic echocardiography.

Using LVM_CMR_ as the reference, we examined the correlation of LVMWT_TTE_ and LVM_TTE-DB_, which are commonly used indices for assessing the severity of LV hypertrophy in conventional TTE analysis. The correlations are shown in **Figure 3**. Although LVM_TTE-DB_ showed better correlations with LVM_CMR_ compared to LVMWT_TTE_, there remained a significant difference (mean difference 106 [90 to 123]; p for difference <0.001). In contrast, unlike LVM_TTE-DB_, which fails to account for asymmetric hypertrophy, LVM_SMART_ estimates LVM by delineating the epicardial and endocardial borders of the LV wall from the apical views. As a result, it not only demonstrated improved correlation (PCC 0.843 [0.779 to 0.890], p<0.001) but also showed excellent agreement with LVM_CMR_ (mean difference, 1 [-11 to 13], p for difference = 0.903).

**Figure 3.**
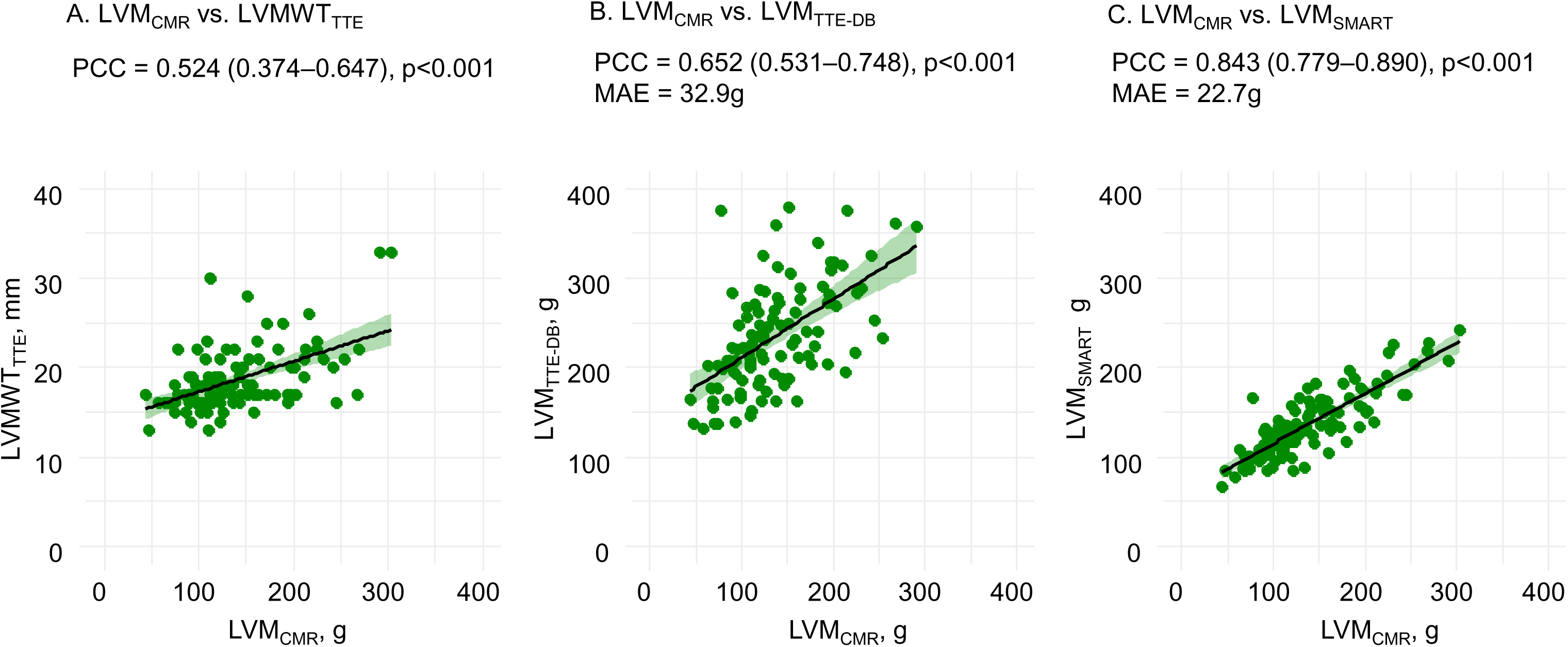
Concordance of SMART and manual measurement to CMR for LVM Compared to LVMWT (LVMWT_TTE_) (A) and dimension-based LVM (LVM_TTE-DB_) (B) measured by TTE, SMART-based LVM (LVM_SMART_) (C) showed stronger concordance to CMR-based LVM (LVM_CMR_). Abbreviations as in Figure 1 and 2: DB, dimension-based methods; LVM, left ventricular mass; LVMWT, left ventricular maximal wall thickness.

We further performed a subgroup analysis stratified by image quality (excellent, n = 22; good, n = 46; fair, n = 43) to assess the performance of SMART under varying imaging conditions. LVGLS_SMART_ maintained strong concordance with LVGLS_TTE_EchoPAC_ across all subgroups, with the highest concordance observed in excellent-quality images (PCC 0.932) and acceptable performance even in fair-quality images (PCC 0.795) (**Supplemental Results 1, 2**). Similarly, LVM_SMART_ consistently demonstrated strong agreement with LVM_CMR_, with PCCs of 0.874, 0.843, and 0.843 for excellent-, good-, and fair-quality images, respectively (**Supplemental Results 3, 4**).

In addition to image quality, we further assessed the performance of SMART according to HCM morphological subtypes. For LVGLS measurement, LVGLS_SMART_ showed good concordance with LVGLS_TTE-EchoPAC_, across all subtypes, with PCCs of 0.808, 0.814, and 0.745 for septal, mixed/diffuse, and apical HCM, respectively (**Supplemental Result 5**).

For LVM measurement, LVM_SMART_ consistently demonstrated strong agreement with LVM_CMR_ across subtypes, with PCCs of 0.821, 0.818, and 0.768, respectively (**Supplemental Result 6**). Representative cases demonstrating the application of SMART across different HCM subtypes are shown in **Figure 4**, alongside corresponding LVGLS_TTE-EchoPAC_ and LVM_CMR_ values.

**Figure 4.**
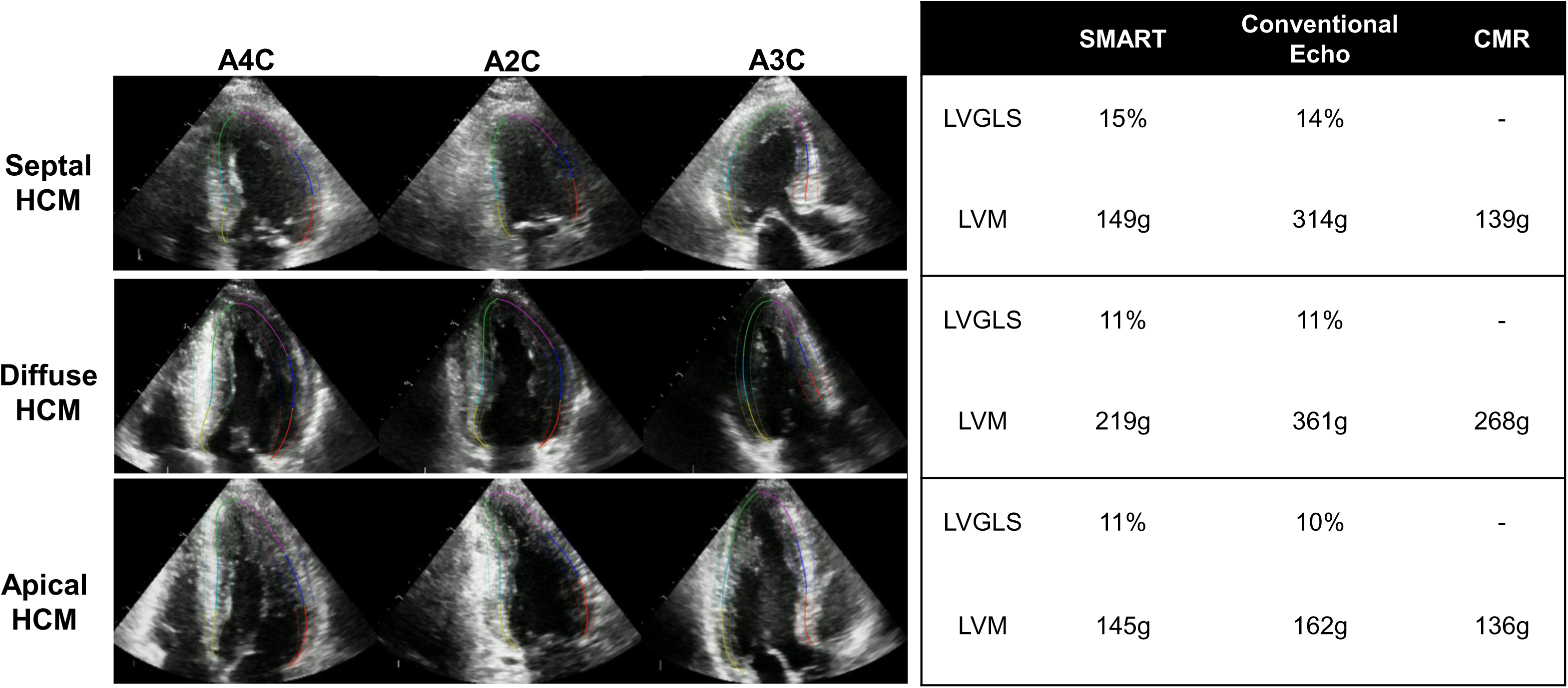
Representative cases for SMART application in HCM patients Representative cases with different HCM subtypes demonstrated that SMART provided consistent assessments of LVGLS compared to manual measurements, as well as LVM values closely aligned with CMR-derived estimates. For “Conventional Echo” values, LVM was calculated using the dimension-based method, which may overestimate or underestimate LVM in cases with asymmetric hypertrophy. Abbreviations as in Figure 1, 2, and 3: A2C, apical 2-chamber view; A3C, apical 3-chamber view; A4C, apical 4-chamber view.

### Predictive Value of SMART based LVGLS and LVM for Extensive LGE

We performed ROC curve analysis to assess whether extensive LGE (>15% of LVM) observed on CMR could be predictive using LVGLS_SMART_. The AUC was 0.72 (0.58 to 0.86), comparable to LVGLS_TTE-EchoPAC_ (AUC 0.75 [0.64 to 0.86], p for difference =0.708) and showing no significant difference compared to LVGLS_CMR_ (AUC 0.78 [0.68 to 0.88], p for difference =0.212) (**Figure 5**). Similarly, we evaluated whether indices used for assessing LV hypertrophy could predict extensive LGE. Conventional TTE-based metrics showed limited predictive ability (AUC 0.59 [0.43 to 0.74] for LVMWT_TTE_; AUC 0.57 [0.39 to 0.74] for LVM_TTE-DB_).

**Figure 5.**
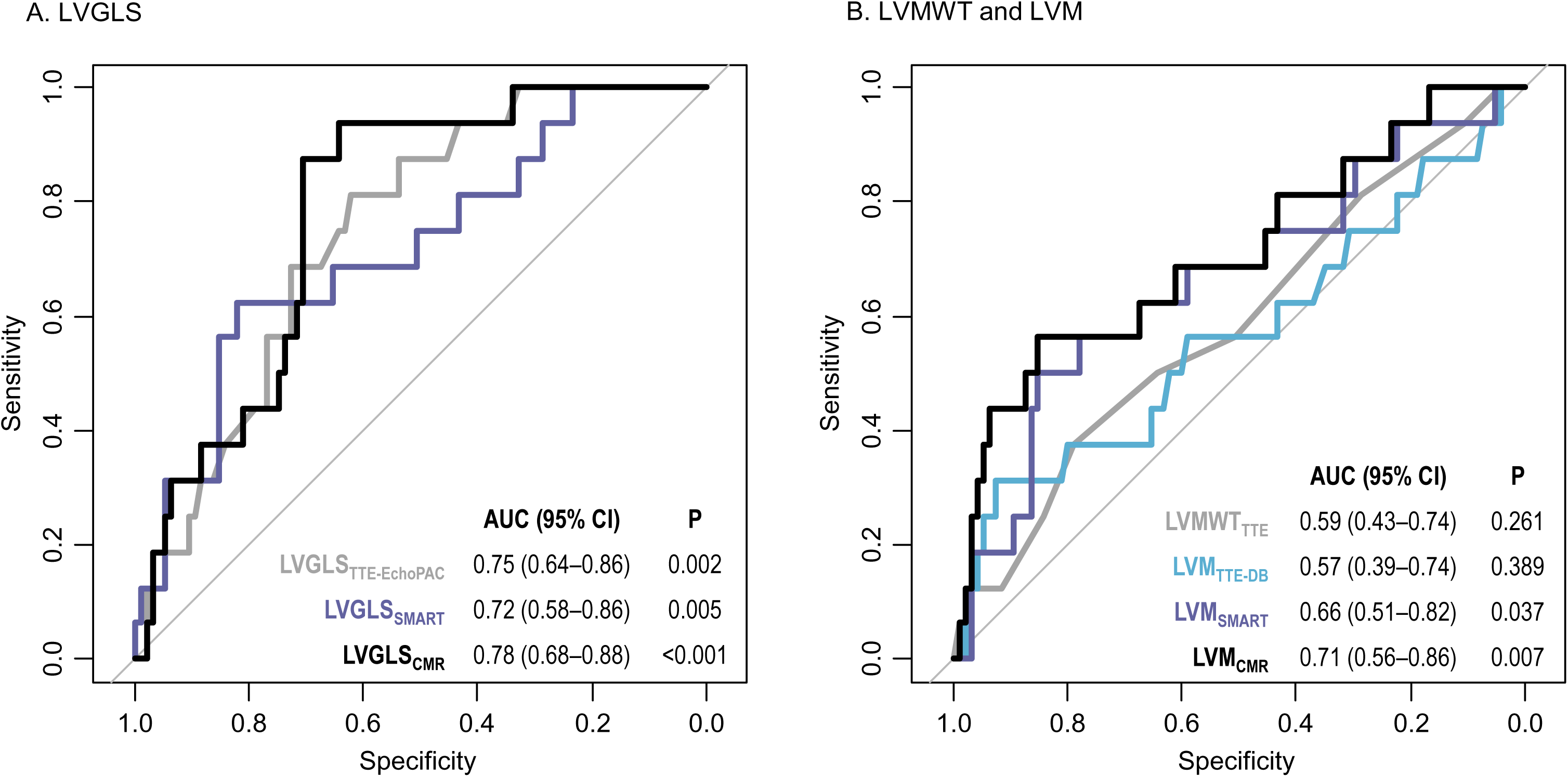
Predictive value of SMART-based LVGLS and LVM for extensive LGE Compared to conventional measurements, SMART-based LVGLS (LVGLS_SMART_) showed comparable performance for predicting extensive LGE (A). SMART-based LVM (LVM_SMART_) outperformed LVMWT (LVMWT_TTE_) and dimension-based LVM (LVM_TTE-DB_) for predicting extensive LGE (B). Abbreviations as in Figure 1-3: AUC, area under the receiver operating characteristic curves; CI, confidence interval; LGE, late gadolinium enhancement.

However, when LVM was measured using the SMART technique, the AUC increased to 0.66 (0.51 to 0.82), which was significantly higher than LVMWT_TTE_ (p for difference = 0.045) and LVM_TTE-DB_ (p for difference = 0.036), but comparable to LVM_CMR_ with no significant difference observed (AUC 0.71 [0.56 to 0.86], p for difference =0.130).

### Outcome Risk Stratification Based on Extensive LGE Predicted by Auto-Measured

#### LVGLS and LVM

During a median follow-up of 7.4 years (5.7-8.8 years), a total of 28 patients experienced the composite outcomes. Specifically, no deaths were observed, while heart failure hospitalization occurred in 2 patients, new-onset atrial fibrillation in 14 patients, and ICD implantation in 15 patients.

The optimal cutoff values for extensive LGE were identified as 11% for LVGLS_SMART_ and 166 g for LVM_SMART_. Patients categorized as high-risk for extensive LGE based on LVGLS_SMART_ (≤11%) had a significantly higher risk of adverse clinical outcomes, with an adjusted HR of 6.02 (2.21 to 16.41) (**Figure 6**). Similarly, those classified as high-risk based on LVM_SMART_ (≥166 g) also demonstrated an increased risk of clinical outcomes, with an adjusted HR of 3.97 (1.50 to 10.47).

**Figure 6.**
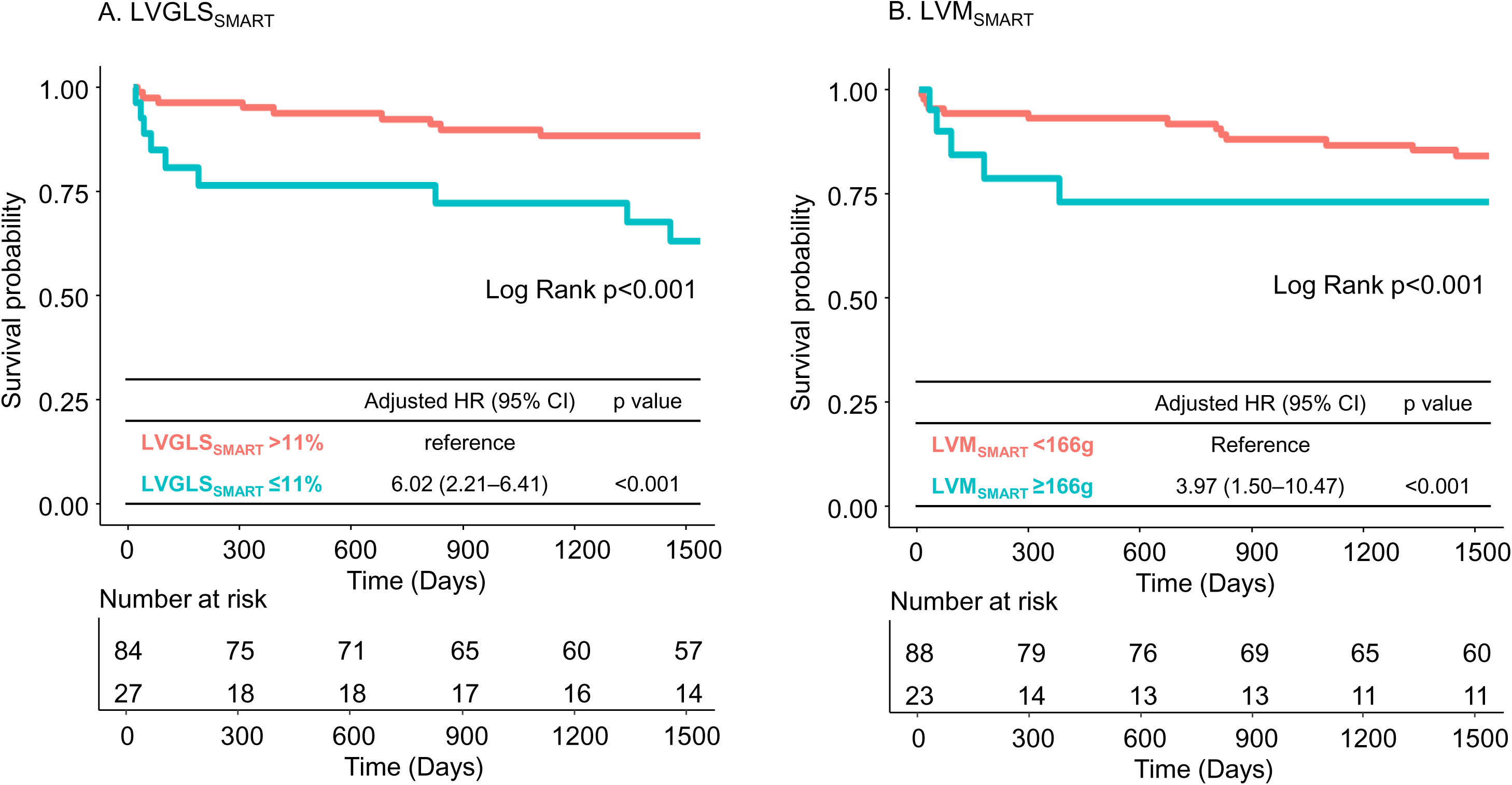
Outcome Risk Stratification Based on SMART-based LVGLS and LVM. Patients identified as high-risk for extensive LGE based on SMART-based LVGLS (LVGLS_SMART_ ≤11%) (A) and LVM (LVM_SMART_ ≥166g) (B) had a significantly higher risk of adverse clinical outcomes. Abbreviations as in Figure 1-4: HR, hazard ratio

## DISCUSSION

In this study, we introduce the SMART technique, a novel approach that refines motion tracking by incorporating LV myocardial segmentation throughout the cardiac cycle. We demonstrated that the SMART technique is feasible and effectively segments LV walls in HCM patients, enabling accurate assessment of LVM relative to CMR and comparable measurement of LVGLS to conventional manual methods. For predicting extensive LGE, LVGLS_SMART_ showed predictive value comparable to LVGLS measured by well-stablished systems, while LVM_SMART_ demonstrated superior predictive value compared to conventional TTE measures such as LVMWT_TTE_ or LVM_TTE-DB_. Additionally, we confirmed the independent prognostic value of both LVGLS_SMART_ and LVM_SMART_.

Conventional speckle tracking system often relies on semi-automated, operator-defined endocardial border delineation, whether tracking the endocardium or mid-myocardium. While effective for assessing global LV motion, this approach does not precisely define both endocardial and epicardial borders to establish an ROI for tracking. Recently, AI-based automatic LV strain analysis has been introduced, enabling automatic segmentation and tracking of the LV wall [9, 19, 25]. Salte et al. demonstrated the feasibility and accuracy of automated LVGLS measurement in 200 patients with various cardiovascular pathologies and levels of LV function, using DL-based LV wall segmentation and mid-wall tracking [11]. They later reported improved precision and reproducibility with this approach [25]. Another study involving 870 COVID-19 patients showed that AI-derived LVEF and LVGLS, based on LV endocardial border tracing, had superior accuracy in predicting mortality compared to manual measurement [26]. We recently presented a system that automatically segments the LV wall to define and track the mid-wall, enabling accurate LVEF and LVGLS measurement [19]. In revascularized ST-segment elevation myocardial infarction patients, our system accurately measured LVEF and LVGLS compared to manual measurements and demonstrated comparable prognostic value. However, most of these studies focused on patients with varying levels of LV dysfunction and did not specifically address LV hypertrophy. The ability of DL-based techniques to accurately delineate the endocardial and epicardial borders and track LV myocardial motion in HCM patients, characterized by severe and asymmetric LV hypertrophy, remains insufficiently validated.

Accurate tracking of LV wall motion requires precise and automated segmentation, which is particularly challenging in TTE due to lower tissue contrast, susceptibility to artifacts, and limited resolution compared to CMR. This aligns with the well-documented challenge of accurately quantifying LV wall hypertrophy in HCM using TTE. For instance, prior studies have reported discrepancies between LVMWT measured by CMR and TTE [27, 28]. Moreover, conventional TTE-derived LVM, which relies on geometric assumptions, often fails to capture asymmetric hypertrophy patterns, such as those seen in septal or apical HCM, as demonstrated in the present study. In contrast, the SMART technique addresses these limitations by integrating accurate LV wall segmentation from A4C, A2C, and A3C views to refine speckle tracking. This approach provides a more precise representation of LV structure, even in cases with asymmetric geometry, resulting in LVM_SMART_ showing high concordance with LVM_CMR_. Given the critical prognostic value of LVM in HCM, the ability to automatically and accurately measure LVM using TTE carries significant clinical implications. In our study, LVM_SMART_, unlike LVMWT_TTE_ or LVM_TTE-DB_, successfully predicted extensive LGE and demonstrated independent prognostic value. This capability may enable more accurate risk stratification and reduce reliance on CMR in routine practice, particularly for patients with asymmetric hypertrophy or limited access to advanced imaging modalities.

Even with fully automated LV segmentation without manual intervention, the SMART technique demonstrated high concordance in LVGLS measurement. By performing segmentation across all frames rather than being limited to end-systole, SMART refines motion tracking and enables reliable and consistent LVGLS quantification. As a result, SMART-derived LVGLS showed strong agreement with expert manual LVGLS measurement obtained using well-established systems. Although concordance with CMR was lower, this was expected due to differences in imaging modalities and examination dates. LVGLS_CMR_ was measured on the cine-CMR conducted on a separate day with non-identical imaging planes, whereas TTE-derived LVGLS measurements were obtained from the same views of the same echocardiographic exam using different methods. Despite these differences, LVGLS_SMART_ demonstrated a predictive performance for extensive LGE comparable to LVGLS_CMR,_ and also showed independent prognostic value. Given the reproducibility and efficiency of SMART-derived GLS, this parameter may serve as a practical tool for longitudinal monitoring of myocardial function, particularly in the context of emerging pharmacologic therapies such as Mavacamten.

Beyond its technical feasibility, the SMART technique is designed to be adaptable to various clinical environments. It operates efficiently on standard workstations equipped with a central processing unit (CPU), without requiring a graphics processing unit (GPU) for acceleration. Complete analysis can typically be achieved in under 10 seconds using CPU-based systems, and in under 5 seconds when GPU acceleration is available. Multiple deployment models are supported, including standalone workstations, integration into echocardiographic imaging systems, PACS-based analysis, and EMR-linked reporting. Such flexibility may facilitate future integration into diverse clinical workflows. Further studies are warranted to evaluate the real-world clinical performance, scalability, and broader applicability of the SMART technique across different patient populations and institutional settings.

Despite these promising results, several limitations need to be acknowledged. Although this study demonstrated the feasibility of the SMART technique in HCM, the sample size was relatively small. Nevertheless, the study cohort included a unique structure incorporating LV geometric and three different LVGLS parameters derived from both TTE and CMR, allowing for a robust comparison of SMART’s performance against these imaging modalities. The SMART technique demonstrated comparable results to CMR in the assessment of LVGLS and LVM. However, the current findings could not be extrapolated to the point that the SMART technique replaces CMR in HCM management. Nevertheless, given the high clinical utility of TTE as a primary screening tool and for repeated follow-up imaging in HCM patients, the SMART technique not only enhances the accuracy of conventional TTE assessment but may also facilitate the identification of patients at high risk for myocardial fibrosis, thereby contributing to additional risk evaluation through advanced diagnostic modalities. While the SMART technique is fully automated, clinical implementation may benefit from adjustable functionality depending on the integrated environment. An interactive version with optional user control is currently under development to support broader real-world applications. In addition, manual GLS measurements were performed by a single expert reader. While EchoPAC-based strain analysis has been shown to be reproducible in prior literature, the use of a single reader may still limit generalizability and should be acknowledged as a methodological constraint. Another limitation is that this study did not directly compare the SMART technique with other AI-based LVGLS or LVM quantification tools, as methodological differences and technical variability across systems would require a separate standardized comparative study. Furthermore, although the SMART algorithm was technically validated in diverse populations—including an American cohort and cross-modality datasets— the present clinical validation was performed exclusively in an East Asian HCM population. Future studies involving multi-ethnic and multi-center cohorts are warranted to confirm the generalizability and clinical applicability of the SMART technique across broader populations and cardiomyopathy subtypes.

## CONCLUSION

The SMART technique is an AI-based novel technique that combines LV speckle tracking with myocardial segmentation in a hybrid approach based on TTE. In patients with HCM, characterized by LV hypertrophy with asymmetric geometry, the SMART technique allows for comparable evaluation of LVGLS and more accurate assessment of LVM compared to conventional measurements in TTE. Furthermore, SMART-derived LVGLS and LVM demonstrated utility in predicting myocardial fibrosis and clinical outcomes, supporting the diagnostic and prognostic feasibility of this novel technique in the management of HCM.

## Author Contributions

J.P. and Y.E.Y. contributed to the conception and design of the study.

J.P., H.M.C., I.C.H., E.J.C., and G.Y.C. were responsible for clinical data acquisition, image curation, expert manual measurements, and independent data analysis and interpretation.

Y.J., T.J., J.J., and S.A.L. (Ontact Health Inc.) contributed to the technical development, including the SMART algorithm.

Y.E.Y. and H.J.C. participated in manuscript drafting and critical revision.

All authors reviewed and approved the final version of the manuscript.

## Data Availability

All data produced in the present study are available upon reasonable request to the authors

## Acknowledgments

This work was supported by a grant from the Institute of Information & communications Technology Planning & Evaluation (IITP) funded by the Korea government (Ministry of Science and ICT) (No.2022000972, Development of a Flexible Mobile Healthcare Software Platform Using 5G MEC); and the Medical AI Clinic Program through the NIPA funded by the MSIT. (Grant No.: H0904-24-1002).

## Conflict of Interest Statement

Y.E.Y., Y.J., T.J., J.J., and S.A.L. are currently affiliated with Ontact Health, Inc. Y.J., and S.A.L are co-inventors on a patent related to this work filed by Ontact Health (Method For Providing Information On Strain Quantification And Device For Providing Information On Strain Quantification Using The Same). Y.E.Y., and H.J.C. holds stock in Ontact Health, Inc. The other authors have no conflicts of interest to declare.

## Data Sharing Statement

Data cannot be made publicly available due to ethical restrictions set by the IRB of the study institution; i.e., public availability would compromise patient confidentiality and participant privacy. Please contact the corresponding author to request the minimal anonymised dataset. Researchers with additional inquiries about the deep learning model presented in this study are also encouraged to reach out to the corresponding author.

## Supplemental Methods

**Supplemental Method 1. Dataset for the Development of the SMART Technique**

**Supplemental Methods 2. CMR Acquisition and Analysis**

**Supplemental Method 3. Performance of Automatic View Classification on the HCM Dataset**

**Supplemental Method 4. Technical Detail of the SMART Technique**

**Supplemental Method 5. Extended Technical Details: SMART Processing Time and Workflow**

**Supplemental Method 6. In-House Validation of SMART-based LVGLS Measurements**

**Supplemental Method 7. Comparison of SMART Technique with Conventional TTE and CMR Measurement in LV volumes and LVM**

## Supplemental Results

**Supplemental Result 1. Subgroup analysis of LVGLS according to image quality**

**Supplemental Result 2. Bland–Altman plots of LVGLS according to image quality**

**Supplemental Result 3. Subgroup analysis of LVM according to image quality**

**Supplemental Result 4. Bland–Altman plots of LVM according to image quality**

**Supplemental Result 5. Subgroup analysis for LVGLS according to HCM phenotype**

**Supplemental Result 6. Subgroup analysis for LVGLS according to HCM phenotype**

## Notes

### Competing Interest Statement

H.J. Chang holds stock in Ontact Health, Inc. Y. Jang, Y.E. Yoon, J. Jeon, T. Jung, Y. Hong, and S.A. Lee are currently affiliated with Ontact Health, Inc. The remaining authors report no conflicts of interest.

### Summary of Updates

This version of the manuscript includes the following updates: methods section revised with additional technical details; supplemental file updated; author list and affiliations revised; and Reference 15 updated with full citation details.

## Reference

1. Ommen SR, Ho CY, Asif IM, t al. 2024 AHA/ACC/AMSSM/HRS/PACES/SCMR Guideline for the Management of Hypertrophic Cardiomyopathy: A Report of the American Heart Association/American College of Cardiology Joint Committee on Clinical Practice Guidelines. Circulation. 2024;149(23):e1239–e311.

2. Mandeş L, Roşca M, Ciupercă D, et al. The role of echocardiography for diagnosis and prognostic stratification in hypertrophic cardiomyopathy. J Echocardiogr. 2020;18(3):137–48.

3. Lang RM, Badano LP, Mor-Avi V, et al. Recommendations for cardiac chamber quantification by echocardiography in adults: an update from the American Society of Echocardiography and the European Association of Cardiovascular Imaging. J Am Soc Echocardiogr. 2015;28(1):1–39 e14.

4. Devereux RB, Alonso DR, Lutas EM, et al. Echocardiographic assessment of left ventricular hypertrophy: comparison to necropsy findings. Am J Cardiol. 1986;57(6):450–8.

5. Kristensen CB, Myhr KA, Grund FF, et al. A new method to quantify left ventricular mass by 2D echocardiography. Sci Rep. 2022;12(1):9980.

6. Dohy Z, Szabo L, Toth A, et al. Prognostic significance of cardiac magnetic resonance-based markers in patients with hypertrophic cardiomyopathy. Int J Cardiovasc Imaging. 2021;37(6):2027–36.

7. Olivotto I, Maron MS, Autore C, et al. Assessment and significance of left ventricular mass by cardiovascular magnetic resonance in hypertrophic cardiomyopathy. J Am Coll Cardiol. 2008;52(7):559–66.

8. Zhang J, Gajjala S, Agrawal P, t al. Fully Automated Echocardiogram Interpretation in Clinical Practice. Circulation. 2018;138(16):1623–35.

9. Yoon YE, Kim S, Chang HJ. Artificial Intelligence and Echocardiography. J Cardiovasc Imaging. 2021;29(3):193–204.

10. Zhou J, Du M, Chang S, et al. Artificial intelligence in echocardiography: detection, functional evaluation, and disease diagnosis. Cardiovasc Ultrasound. 2021;19(1):29.

11. Salte IM, Ostvik A, Smistad E, et al. Artificial Intelligence for Automatic Measurement of Left Ventricular Strain in Echocardiography. JACC Cardiovasc Imaging. 2021;14(10):1918– 28.

12. Kwan AC, Chang EW, Jain I, et al. Deep Learning-Derived Myocardial Strain. JACC Cardiovasc Imaging. 2024;17(7):715–25.

13. MA Azad, A Chernyshov, J Nyberg, et al. EchoTracker: Advancing Myocardial Point Tracking in Echocardiography. arXiv [cs.CV; 2024]. Available from: https://arxiv.org/abs/2405.08587.

14. A Chernyshov, J Nyberg, V Holmstrøm, et al. Low Complexity Point Tracking of the Myocardium in 2D Echocardiography. arXiv [eess.IV; 2025]. Available from: https://arxiv.org/abs/2503.10431

15. Jeon J, Ha S, Yoon YE, et al. Echocardiographic view classification with integrated out-of-distribution detection for enhanced automatic echocardiographic analysis. arXiv [eess.SP; 2023]. Available from: https://arxiv.org/abs/2308.16483.

16. Jeon J, Kim J, Jang Y, et al. A Unified Approach for Comprehensive Analysis of Various Spectral and Tissue Doppler Echocardiography. arXiv [eess.IV; 2023]. Available from: https://arxiv.org/abs/2311.08439.

17. Jeong D, Jung S, Yoon YE, et al. Artificial intelligence-enhanced automation for M-mode echocardiographic analysis: ensuring fully automated, reliable, and reproducible measurements. Int J Cardiovasc Imaging. 2024;40(6):1245–56.

18. Park J, Jeon J, Yoon YE, et al. Artificial intelligence-enhanced automation of left ventricular diastolic assessment: a pilot study for feasibility, diagnostic validation, and outcome prediction. Cardiovascular Diagnosis and Therapy. 2024;14(3):352–66.

19. Jang Y, Choi H, Yoon YE, et al. An Artificial Intelligence-Based Automated Echocardiographic Analysis: Enhancing Efficiency and Prognostic Evaluation in Patients With Revascularized STEMI. Korean Circ J. 2024;54(11):743–56.

20. Park J, Kim J, Jeon J, et al. Artificial Intelligence-Enhanced Comprehensive Assessment of the Aortic Valve Stenosis Continuum in Echocardiography. medRxiv 2024.07.08.24310123. Available from: 10.1101/2024.07.08.24310123.

21. Agency NIS. Open AI Dataset Project (AI-Hub) [Available from: https://aihub.or.kr/.

22. Park J, Yoon YE, Chun EJ, et al. Endocardial versus whole-myocardial tracking global longitudinal strain analysis in patients with hypertrophic cardiomyopathy: A preliminary comparative study. PLoS One. 2023;18(7):e0288421.

23. Yoon YE, Kang SH, Choi HM, et al. Prediction of infarct size and adverse cardiac outcomes by tissue tracking-cardiac magnetic resonance imaging in ST-segment elevation myocardial infarction. Eur Radiol. 2018;28(8):3454–63.

24. Chan RH, Maron BJ, Olivotto I, et al. Prognostic value of quantitative contrast-enhanced cardiovascular magnetic resonance for the evaluation of sudden death risk in patients with hypertrophic cardiomyopathy. Circulation. 2014;130(6):484–95.

25. Salte IM, Ostvik A, Olaisen SH, et al. Deep Learning for Improved Precision and Reproducibility of Left Ventricular Strain in Echocardiography: A Test-Retest Study. J Am Soc Echocardiogr. 2023;36(7):788–99.

26. Asch FM, Descamps T, Sarwar R, et al. Human versus Artificial Intelligence-Based Echocardiographic Analysis as a Predictor of Outcomes: An Analysis from the World Alliance Societies of Echocardiography COVID Study. J Am Soc Echocardiogr. 2022;35(12):1226–37.e7.

27. Urbano-Moral JA, Gonzalez-Gonzalez AM, Maldonado G, et al. Contrast-Enhanced Echocardiographic Measurement of Left Ventricular Wall Thickness in Hypertrophic Cardiomyopathy: Comparison with Standard Echocardiography and Cardiac Magnetic Resonance. J Am Soc Echocardiogr. 2020;33(9):1106–15.

28. Hindieh W, Weissler-Snir A, Hammer H, et al. Discrepant Measurements of Maximal Left Ventricular Wall Thickness Between Cardiac Magnetic Resonance Imaging and Echocardiography in Patients With Hypertrophic Cardiomyopathy. Circ Cardiovasc Imaging. 2017;10(8):e006309.

