## Supplementary material for "Novel Deep Learning Framework for Simultaneous Assessment of Left Ventricular Mass and Longitudinal Strain: Clinical Feasibility and Validation in Patients with Hypertrophic Cardiomyopathy": Supplmental material

**Supplemental Method 1. Dataset for the Development of the SMART Technique**

We conducted in-house validation of the Segmentation-based Myocardial Advanced Refinement Tracking (SMART) technique across diverse study populations.

1. Left ventricular (LV) global longitudinal strain (LVGLS)

- To validate SMART-based LVGLS, 67 subjects were selected from the in-house dataset, all of whom had the required apical views (A4C, A3C, and A2C) with a frame rate of at least 30 frames per second, and ensuring compatibility with GE EchoPAC software.
- Further validation was conducted in an independent cohort of 217 American patients who underwent TTE examinations at Mayo Clinic in Arizona, USA, and Severance Hospital in Seoul, South Korea.

2. LV Volume and Mass

- To validate the SMART technique-derived tri-plane-based LV volume measurement, 65 subjects with all required apical views (A4C, A3C, and A2C) were selected from the in-house dataset.
- Additionally, to validate both LV volume and LVM measurements, a paired dataset comprising 41 patients with suspected coronary artery disease who underwent both TTE and CMR within 3 months at Seoul National University Bundang Hospital, South Korea, was analyzed.

**Supplemental Method 2. CMR Acquisition and Analysis**

CMR images were acquired using either a 1.5-T system (Intera CV release 10; Philips Healthcare, Amsterdam, Netherlands) or a 3-T system (Philips Ingenia; Best, Netherlands). Imaging was performed under electrocardiographic gating and breath-hold conditions. Steady-state free-precession (SSFP) cine-CMR images were obtained in the horizontal long axis, vertical long axis, and LV outflow tract. A short-axis stack view of the whole LV was obtained for LV volume and mass analysis. The cine-CMR sequence was acquired at 25-30 frames/R-R interval (field of view, 320-370 mm; repetition time/echo time, 3.0-3.6/1.5-1.8ms; flip angle, 45-60°; and slice thickness, 6-8 mm). Late gadolinium enhancement (LGE) images were obtained 10 minutes after intravenous administration of 0.2 mmol/kg of gadodiamide (Omniscan; GE Healthcare, Princeton, NJ, USA) using a phase-sensitive inversion-recovery turbo field echo sequence (repetition/echo time: 4.5–4.6/1.3–1.5ms, flip angle: 20–25°, slice thickness: 8 mm).

All acquired images were processed using CVI42 software (version 5.10; Circle Cardiovascular Imaging, Calgary, Canada) and analyzed by an independent radiologist blinded to clinical and echocardiographic data. LV volumes ($LVEDV_{CMR}$ and $LVESV_{CMR}$), $LVEF_{CMR}$, and $LVM_{CMR}$ were estimated from short-axis cine-CMR. LVGLS analysis was conducted via CMR tissue-tracking ($LVGLS_{CMR}$), which uses a mid-surface curvilinear coordinate system to track myocardial deformation and follows the motion of software-generated myocardial nodes on SSFP cine sequences. Endocardial and epicardial borders were traced in a semi-automated fashion, with manual correction for contours with apparent deviation. Using long-axis cine-images, the whole myocardial LVGLS was calculated throughout the cardiac cycle by the software. As with TTE-derived LVGLS values, the peak negative value (peak systolic strain) was converted to an absolute value, designated as

$LVGLS_{CMR}$. LGE mass was quantified using the full-width at half-maximum method. The total LGE mass was obtained by summing the LGE from all sections, and the relative extent of LGE was expressed as a percentage of total LVM. Extensive LGE was defined as involving >15% of LVM, a threshold associated with an increased risk of sudden cardiac death (SCD).

**Supplemental Method 3. Performance of Automatic View Classification on the HCM Dataset**

The confusion matrix summarizes the performance of the automatic view classification module[1] when applied to the current HCM cohort. The overall classification accuracy was 99.6%. Notably, all echocardiographic views that were actually utilized for SMART analysis—apical four-chamber (A4C), apical two-chamber (A2C), and apical three-chamber (A3C)—were correctly classified without mislabeling.

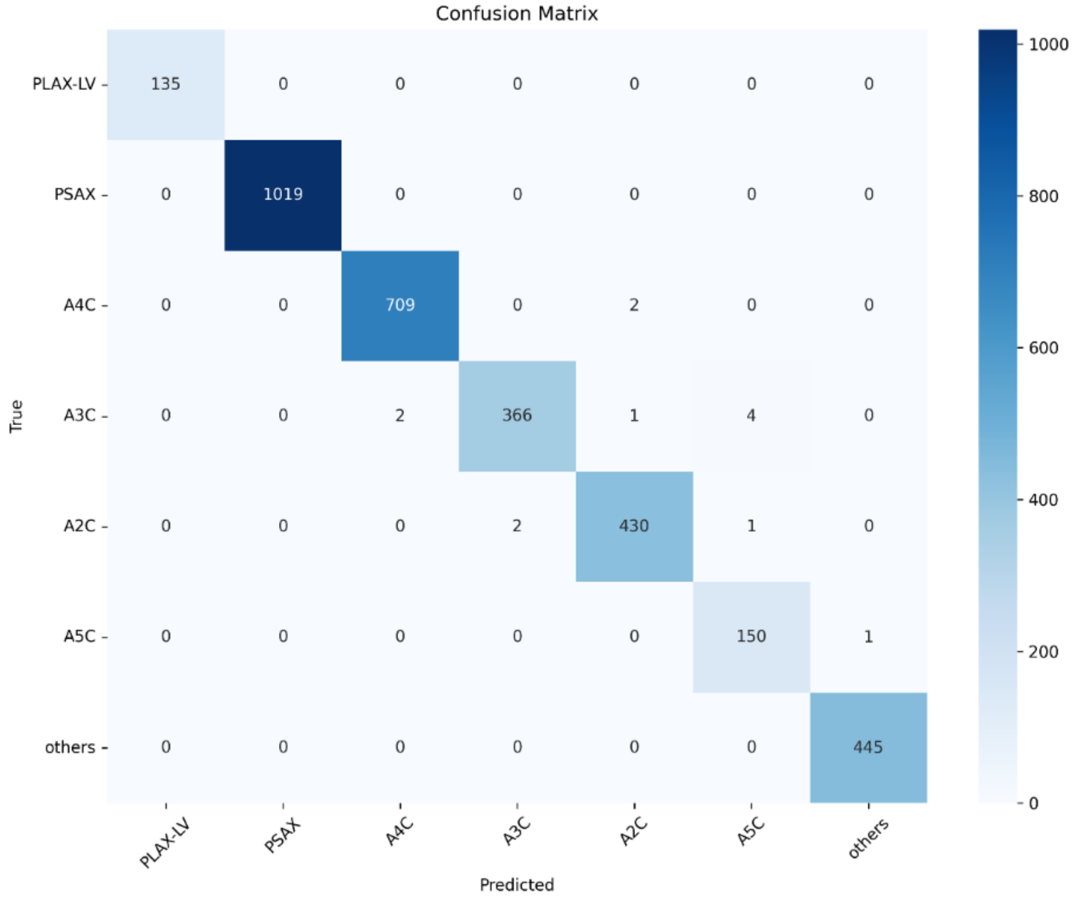

#### Supplemental Method 4. Technical Detail of the SMART Technique

Our AI-based system (Sonix Health Workstation, version 2.0; Ontact Health Inc., Korea) is designed to evaluate key functional metrics of the left ventricle (LV), including volume, mass, and longitudinal strain, through simultaneous segmentation and motion estimation. For a detailed description of the system and its workflow, please refer to our previous study.[2] A summarized diagram of the workflow is provided here for reference.

**Workflow of the fully automated LV quantification system**

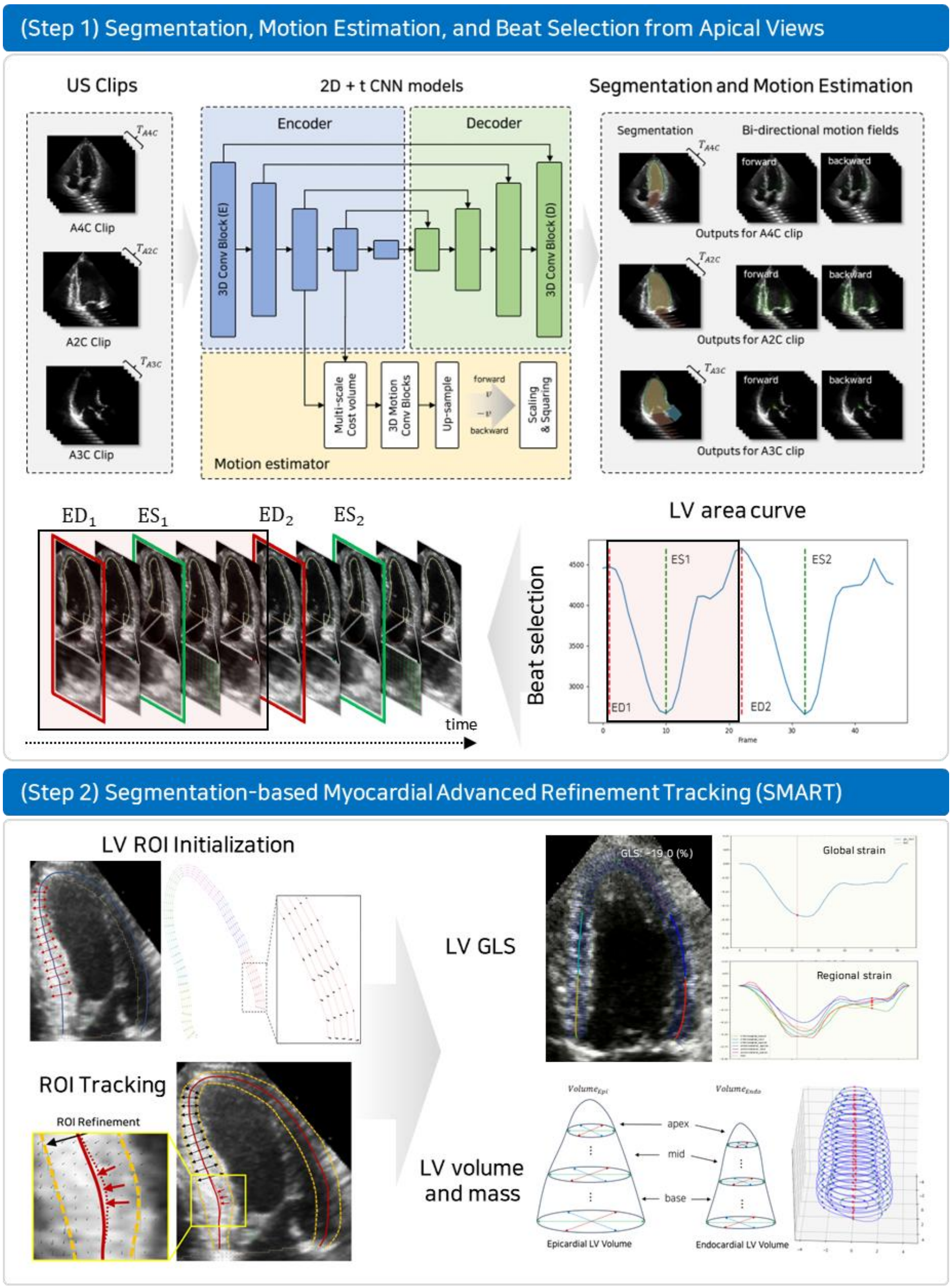

The SMART framework builds upon a 3D CNN architecture to enable precise LV strain quantification. By integrating segmentation and speckle tracking into a unified process, SMART provides robust and accurate LV volume, mass, and strain measurements in 2D echocardiography, establishing its clinical utility for comprehensive LV function assessment. The process begins with LV segmentation across all frames, constructing the LV area curve as the foundation for cardiac cycle analysis. This curve identifies critical cardiac phases, specifically end-diastole (ED) and end-systole (ES).

For longitudinal strain quantification, the LV contour, denoted as $C$, is initially traced at the ES phase and dynamically updated throughout the cardiac cycle using the bi-directional dense motion field $m$ derived from the network. The update process is defined as:

$$C(u,t+1) = C(u,t) + m(C(u,t),t)$$

where $t$ represents time and $u$ signifies the parameterization of the curve within the [0, 1] interval.

The SMART framework employs a two-step refinement process to enhance tracking accuracy. This refinement is applied to the updated region of interest (ROI) curve, with adjustments based on a raster scan along its normal direction. In the first step, the mid-myocardial line within the ROI is refined to better align with the underlying myocardial structure. In the second step, the epicardial and endocardial borders are further adjusted to ensure precise delineation of the LV myocardium. Throughout these steps, the network's segmentation results of the LV myocardium are compared and aligned with the updated curve. This dual refinement process ensures myocardial structure-aware tracking, resulting more precise and reliable measurements.

Finally, the LV strain at time $t$ is calculated as:

$$LV\ strain(t) = \frac{L(t) - L(0)}{L(0)}$$

where $L(t)$ defines the length of the LV contour at time $t$, calculated as $L(t) = \int |C'(u,t)|\, du$. Here, $L(0)$ denotes the length of the LV contour at the ED phase. LV global longitudinal strain (LVGLS) is the average peak strain value obtained from motion tracking in apical 4-, 2- and 3-chamber views.

**Refinement Process in the SMART Framework for Myocardial Structure-Aware Enhanced Tracking**

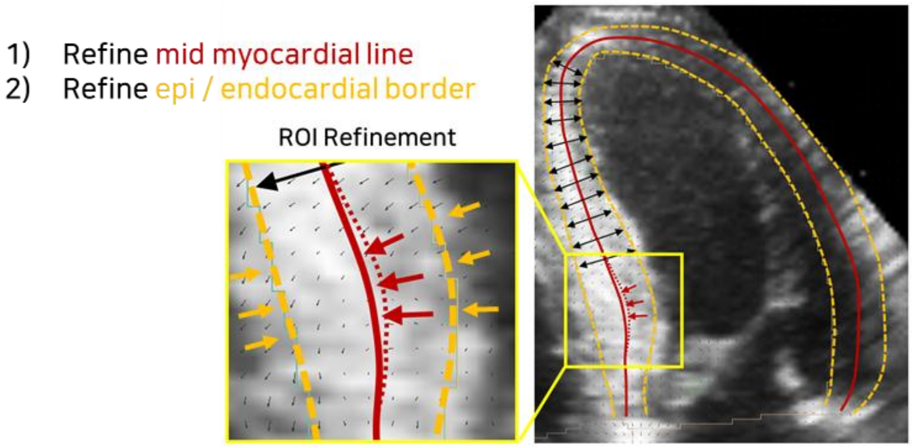


LV volume for the ED and ES phases is calculated using the SMART framework. This framework employs a tri-plane Simpson's method, which offers improved accuracy compared to the conventional biplane approach.[3] The tri-plane method utilizes apical 4-, 2-, and 3-chamber views, enabling a more comprehensive assessment of the LV cavity. Unlike the biplane Simpson's method, the tri-plane approach aligns and maps three planes at 60-degree angles. For each cross-sectional plane, ellipsoid fitting is performed to determine each disk's major and minor axis diameters, allowing for precise volume calculations. The total LV volume ($V_T$) is then calculated by summing the volumes of all individual disks, as expressed in the following equation:

$$V_T = \frac{\pi L}{4n} \sum_{i=1}^{n} a_i * b_i$$

Here, $a_i$ and $b_i$ represent the major and minor axis diameters of the $i^{th}$ disk, $L$ is the length of the LV cavity, and $n$ is the total number of disks. In this study, $n$ is set to 20, a standard value commonly used for this method. At the ED phase, LV mass (LVM) is determined by subtracting the endocardial volume $EDV_{endo}$ from the epicardial volume $EDV_{epi}$. This provides a reliable measure for LVM assessment.

**Quantification of LV Volume and Mass Using Tri-Plane Methods**

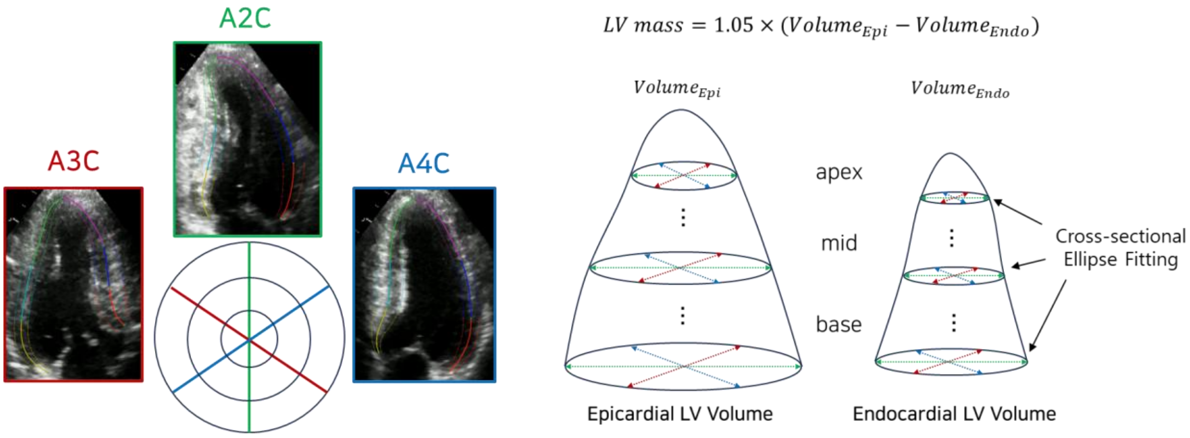

**Supplemental Method 5. Extended Technical Details: SMART Processing Time and Workflow**

To assess the practical feasibility of SMART in real-world clinical environments, processing times were measured using a standard central processing unit (CPU) system (Intel Core i7-12th Gen 12700K, Alder Lake).

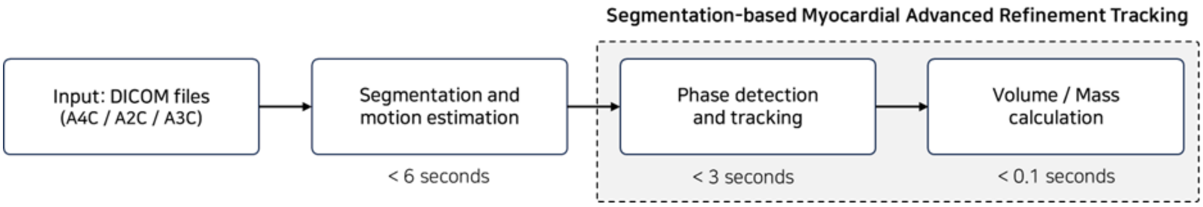


- Segmentation and motion estimation were completed in under 6 seconds.
- Phase detection and motion tracking were completed in under 3 seconds.
- LV volume and mass calculations were completed in under 0.1 seconds per study.

Thus, the full analysis was typically achieved within a few seconds, without requiring GPU acceleration. Processing times are expected to be further reduced if a graphics processing unit (GPU) is available.

**Supplemental Method 6. In-House Validation of SMART-based LVGLS Measurements**

To compare SMART-based LVGLS ($LVGLS_{SMART}$) with manually measured LVGLS using GE EchoPAC software (GE Ultrasound, USA; $LVGLS_{TTE\text{-}EchoPAC}$), TTE images acquired from the GE Echo system were retrospectively collected to construct separate in-house test dataset comprising 67 subjects with all required apical views (A4C, A2C, and A3C). $LVGLS_{SMART}$ demonstrated a strong correlation with $LVGLS_{TTE\text{-}EchoPAC}$ (Pearson Correlation Coefficient = 0.89) and a mean difference of 0.61% (95% CI 0.243-0.974).

**Comparison of SMART and Manual Measurement of LVGLS**

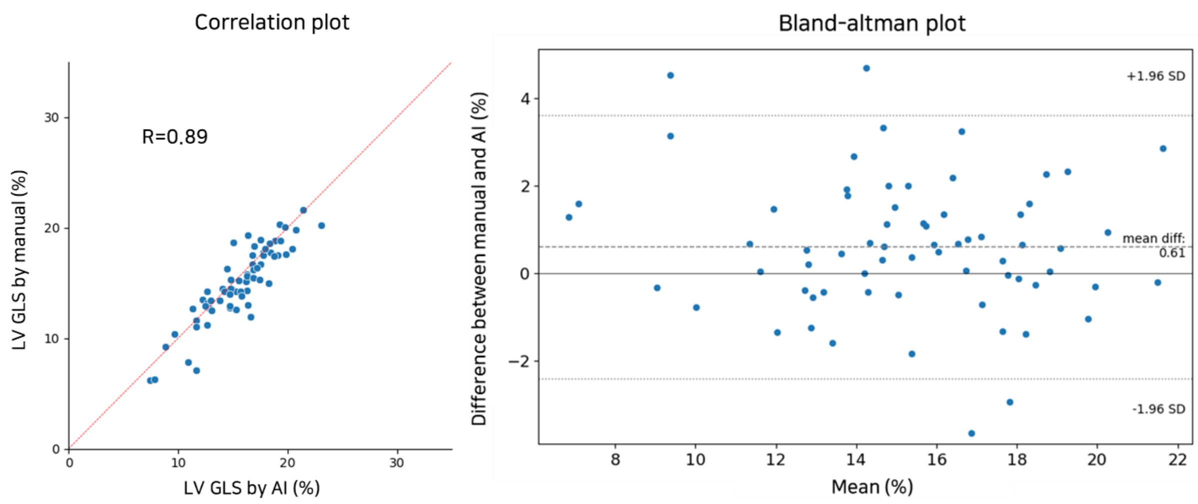

To further evaluate the generalizability of the SMART technique, originally developed using echocardiographic data from Korean patients, we validated it on an additional cohort of 217 American patients. This cohort comprises individuals who underwent transthoracic echocardiographic examinations at Mayo Clinic in Arizona, USA, and Severance Hospital in Seoul, South Korea. In the American population, 87 patients had apical A4C, A2C, and A3C views available for conventional manual measurements using GE EchoPAC software (GE Ultrasound, USA). The SMART technique successfully measured LVGLS in all cases where manual measurements were performed. $LVGLS_{SMART}$ showed strong agreement with $LVGLS_{TTE\text{-}EchoPAC}$, with a Pearson Correlation Coefficients of 0.93, and a mean difference of –0.76% (95% CI -1.083—0.436)

**Comparison of SMART and Manual Measurement for LVGLS in American Cohort**

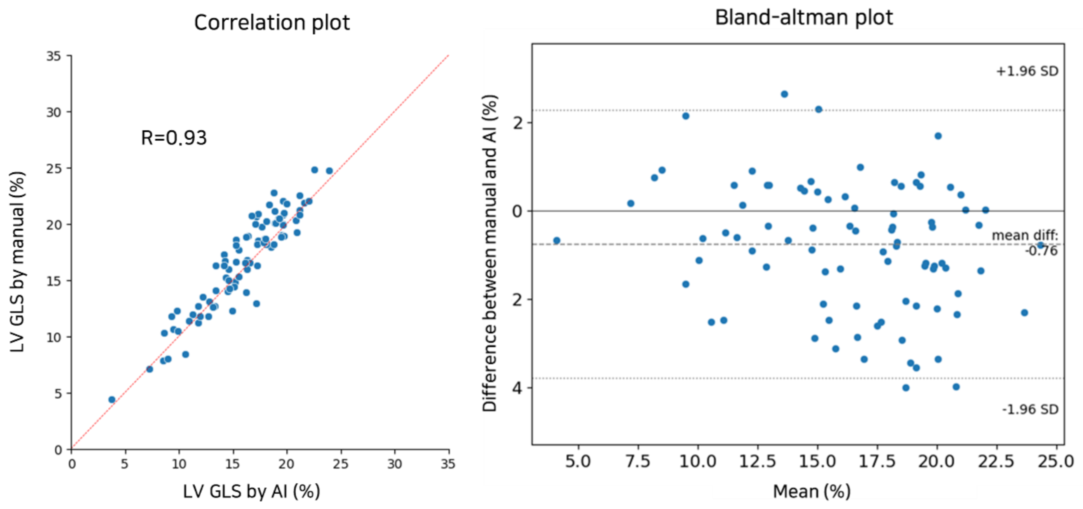

**Supplemental Method 7. Comparison of SMART Technique with Conventional TTE and CMR Measurement in LV volumes and LVM**

We have previously reported the performance of automatic segmentation using the internal test dataset.[2] In this study, the SMART technique was applied to measure LV volumes in 65 subjects with all required apical views (A4C, A3C, and A2C). The automatically measured LV volumes ($LVEDV_{SMART}$ and $LVESV_{SMART}$) were compared with conventional manual measurements obtained using the bi-plane Simpson's method ($LVEDV_{TTE}$ and $LVESV_{TTE}$). Both $LVEDV_{SMART}$ and $LVESV_{SMART}$ showed a strong correlation with $LVEDV_{TTE}$, and $LVESV_{TTE}$ (Pearson Correlation Coefficient = 0.97 and 0.98, respectively), with a mean difference of 3.49mL (95% CI 1.547-5.436) for LVEDV and 2.19mL (95% CI 0.778-3.594) for LVESV.

**Comparison of SMART and Manual Measurement for LV Volumes**

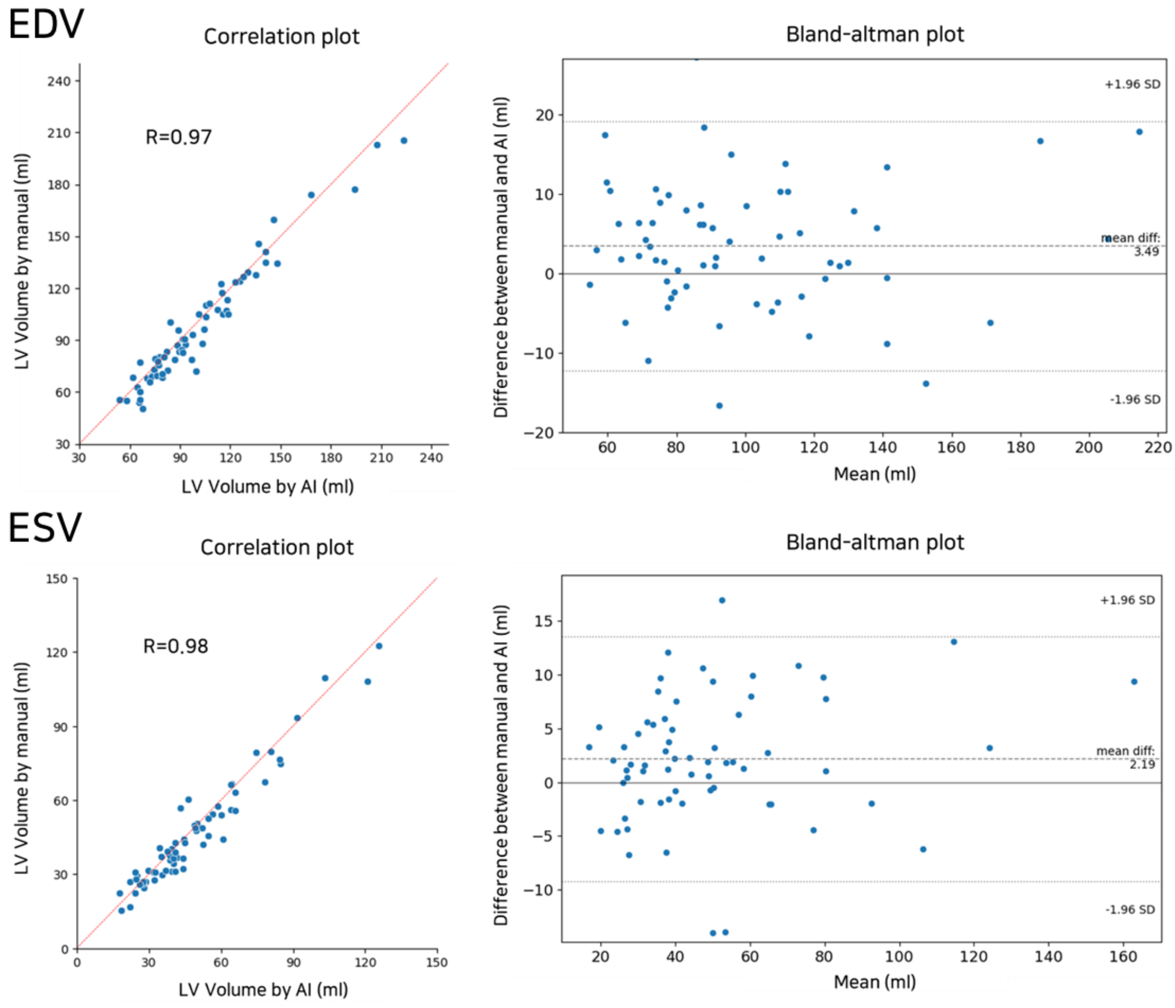

To further evaluate the performance of the SMART technique in measuring both LV volume and LV mass (LVM), its results were compared with cardiac MRI (CMR), the gold standard for such measurements, and conventional manual TTE measurements. This validation was conducted using a paired dataset of 41 patients with suspected coronary artery disease who underwent echocardiography and CMR imaging within three months at Seoul National University Bundang Hospital.

CMR imaging was performed using a 1.5-T system (Intera CV release 10; Philips Healthcare, Amsterdam, Netherlands) or a 3-T system (Philips Ingenia; Best, Netherlands). All scans were conducted under electrocardiographic gating and breath-hold conditions. Steady-state free-precession (SSFP) cine-CMR images were obtained in multiple planes, including the horizontal long axis, vertical long axis, and LV outflow tract. A short-axis stack of the entire LV was used to analyze LV volume and mass precisely.

The SMART-based LV volume ($LVEDV_{SMART}$, and $LVESV_{SMART}$) and mass ($LVM_{SMART}$) were compared with conventional TTE measurement ($LVEDV_{TTE}$, and $LVESV_{TTE}$, and $LVM_{TTE-DB}$) and CMR results ($LVEDV_{CMR}$, $LVESV_{CMR}$, and $LVM_{CMR}$) to evaluate the accuracy and reliability of the SMART technique in this paired dataset.

For $LVEDV_{SMART,}$ a moderate correlation (r=0.54) with $LVEDV_{CMR}$ was observed with a mean difference of -0.26 mL (95% CI: -5.58 to 5.05). In contrast, $LVEDV_{TTE}$ showed a similar correlation (r=0.53) but a more considerable mean difference of -9.45 mL (95% CI: -14.65 to -4.25). Between $LVEDV_{SMART}$ and $LVEDV_{TTE}$, a strong correlation (r=0.89) was observed, with a mean difference of 9.19 mL (95% CI: 6.43 to 11.95). For $LVESV_{SMART}$, comparison with $LVESV_{CMR}$ yielded a correlation of r=0.38 and a mean difference of 7.74 mL (95% CI: 4.71 to 10.78). Similarly, $LVESV_{TTE}$ showed a correlation of r=0.35 and a mean difference of 3.57 mL (95% CI: 0.46 to 6.68). Between $LVESV_{SMART}$ and $LVESV_{TTE}$, a strong correlation (r=0.84) was observed, with a mean difference of 4.17 mL (95% CI: 2.59 to 5.76).

For LVM, $LVM_{SMART}$ demonstrated strong agreement with $LVM_{CMR}$, showing a correlation of r=0.82 and a mean difference of -3.13 g (95% CI: -7.94 to 1.68). By comparison, $LVM_{TTE-DB}$ had a weaker correlation with $LVM_{CMR}$ (r=0.66) and a mean difference of 66.28 g (95% CI: 57.22 to 75.35). Between $LVM_{SMART}$ and $LVM_{TTE-DB}$, a moderate correlation (r=0.64) was observed, with a mean difference of -69.41 g (95% CI: -78.83 to -59.99).

**Correlation Analysis and Bland-Altman for LV Volume and Mass Between SMART, CMR, and TTE**

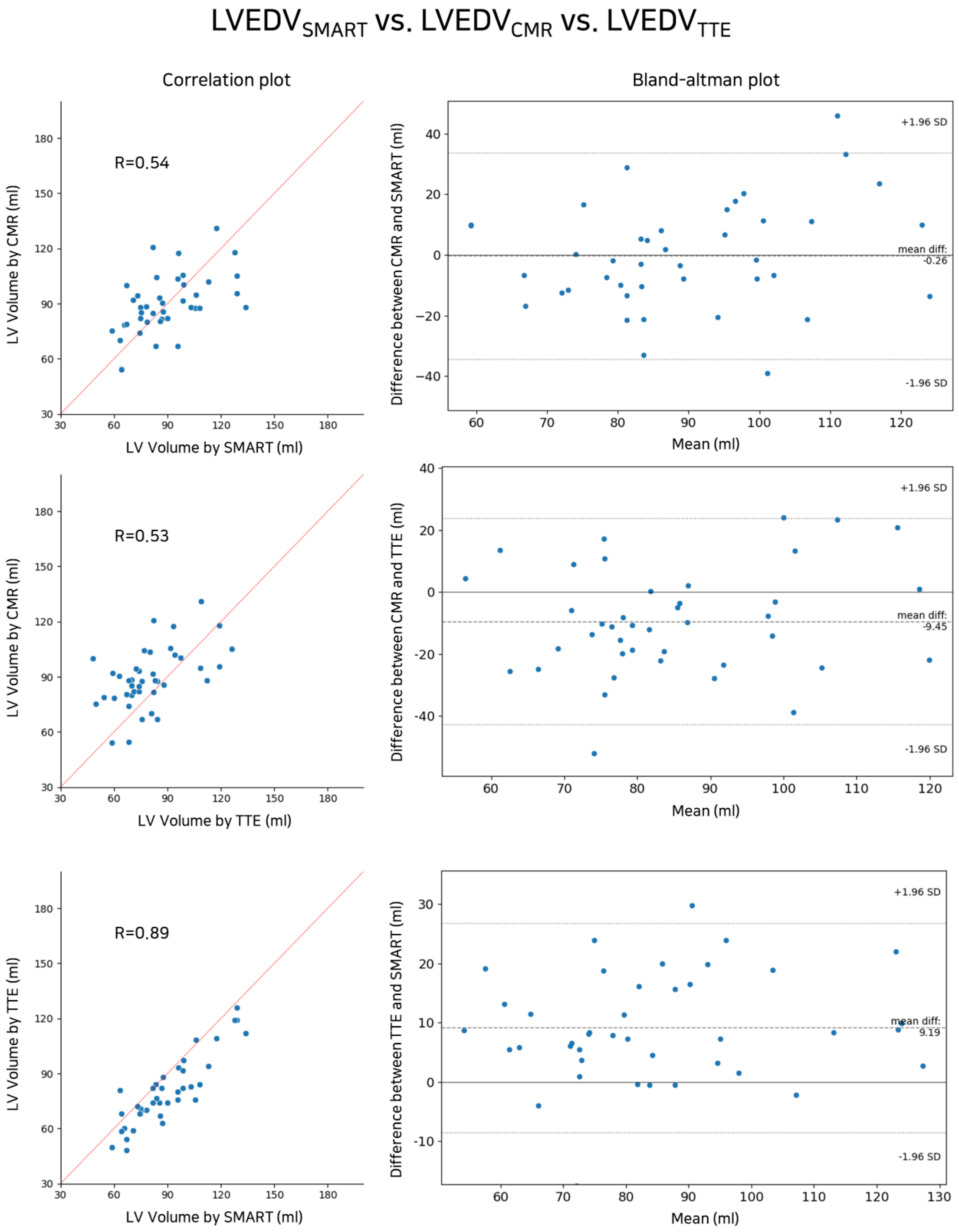

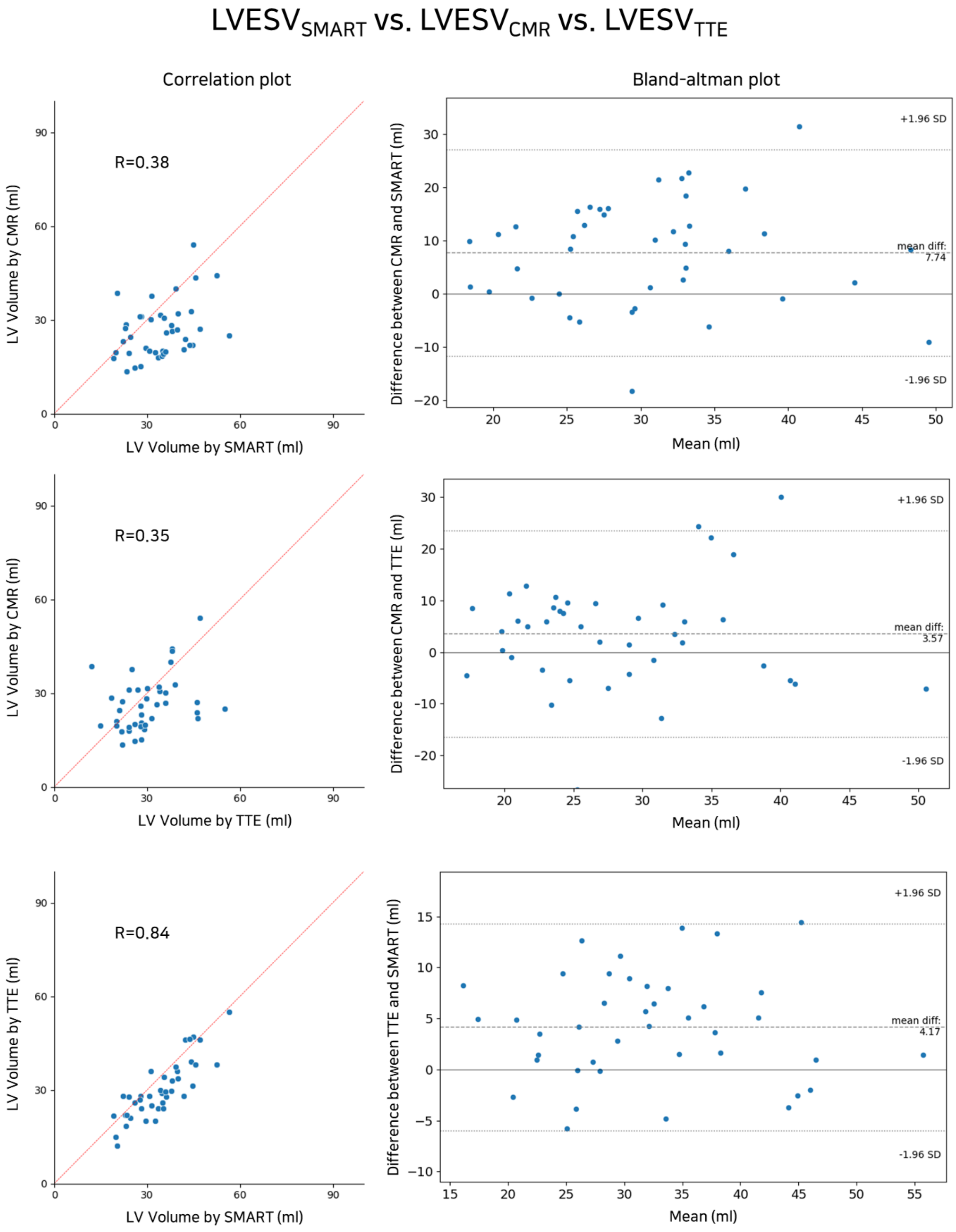

LVESV_SMART vs. LVESV_CMR vs. LVESV_TTE
Correlation plot
Bland-altman plot
R=0.38
LV Volume by CMR (ml)
LV Volume by SMART (ml)
Difference between CMR and SMART (ml)
+1.96 SD
mean diff: 7.74
-1.96 SD
Mean (ml)
R=0.35
LV Volume by CMR (ml)
LV Volume by TTE (ml)
Difference between CMR and TTE (ml)
+1.96 SD
mean diff: 3.57
-1.96 SD
Mean (ml)
R=0.84
LV Volume by TTE (ml)
LV Volume by SMART (ml)
Difference between TTE and SMART (ml)
+1.96 SD
mean diff: 4.17
-1.96 SD
Mean (ml)

### $LVM_{SMART}$ vs. $LVM_{CMR}$ vs. $LVM_{TTE\text{-}DM}$

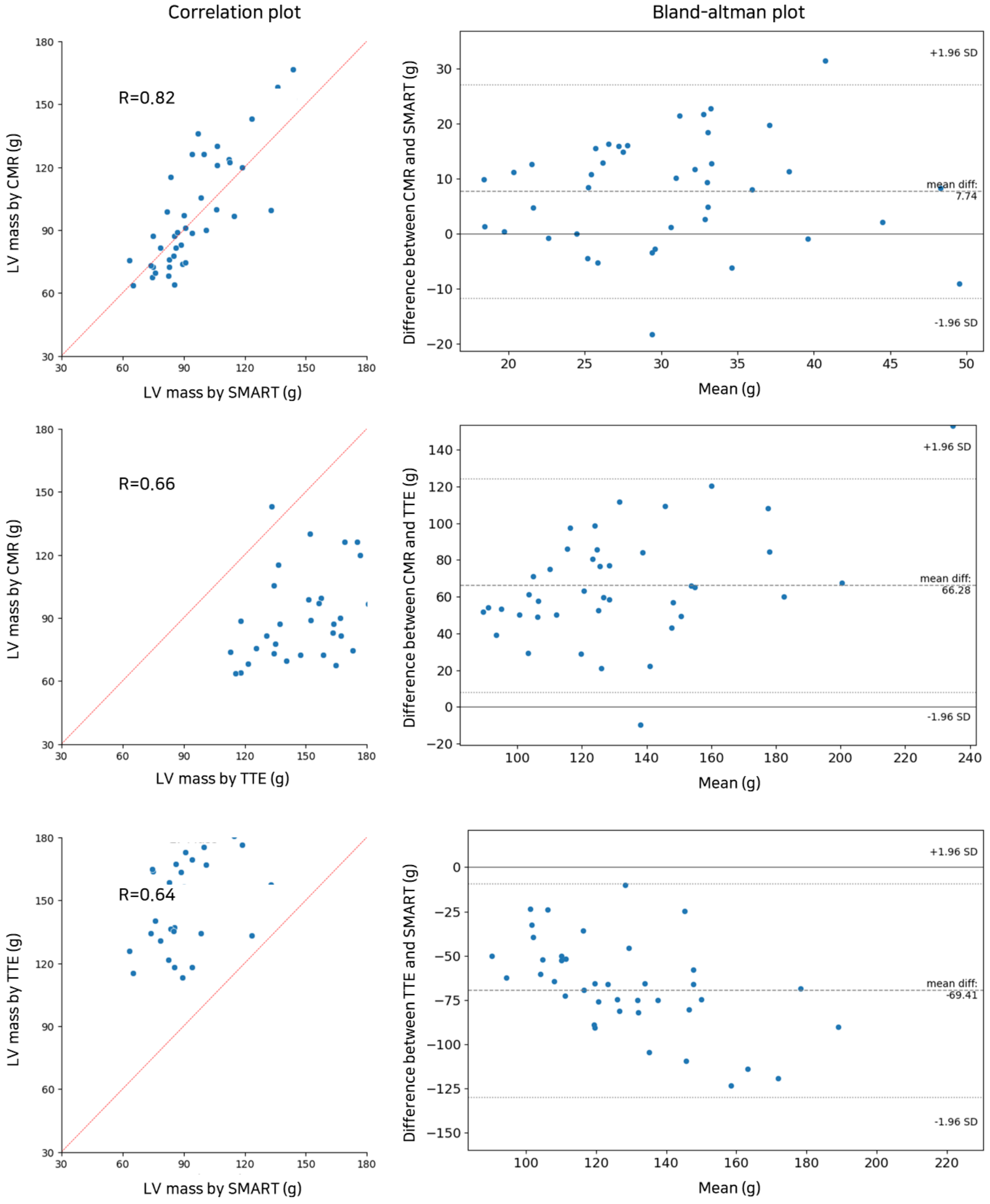

#### Supplemental Result 1. Subgroup analysis of LVGLS according to image quality

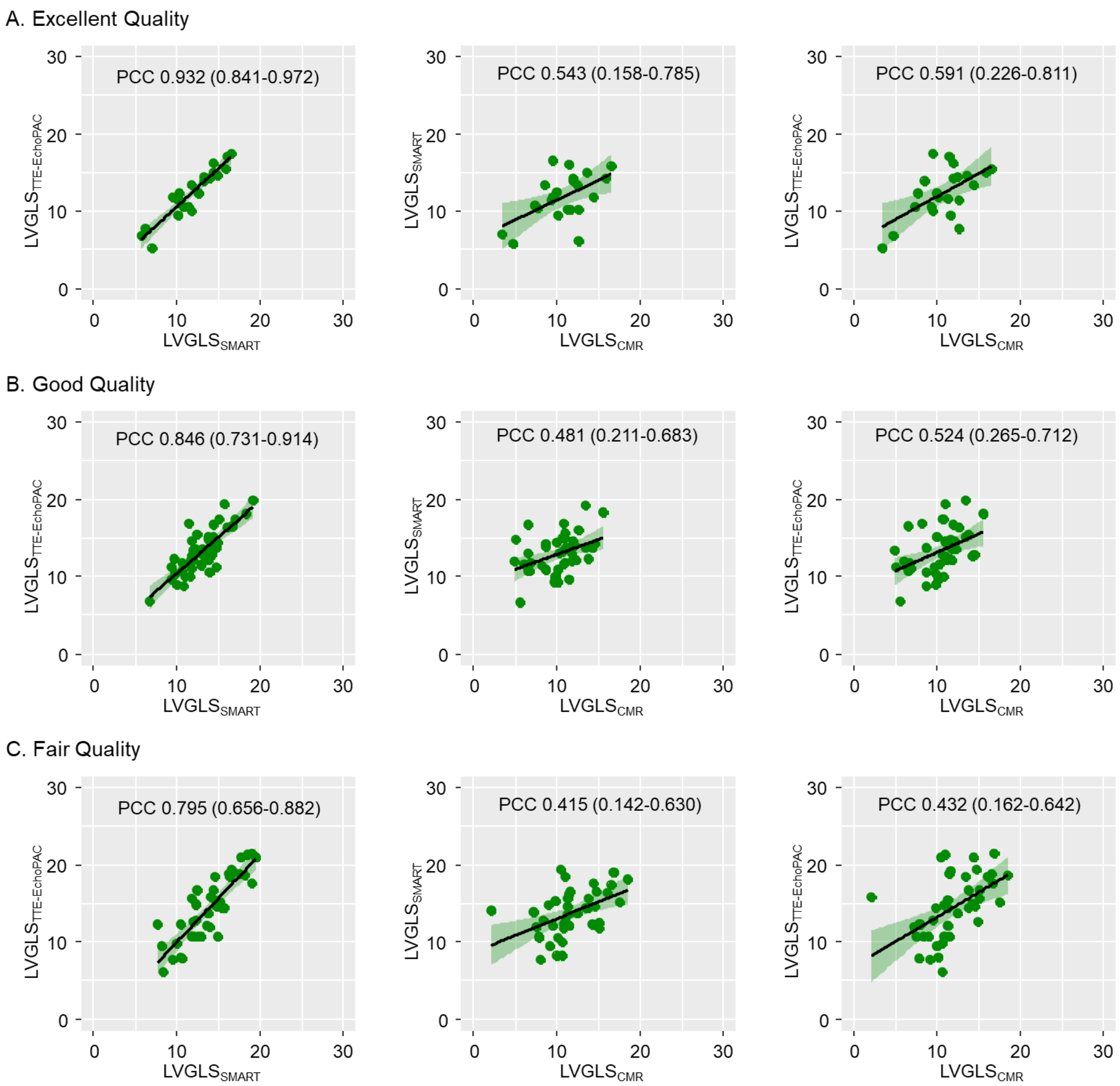

### Supplemental Result 2. Bland–Altman plots of LVGLS according to image quality

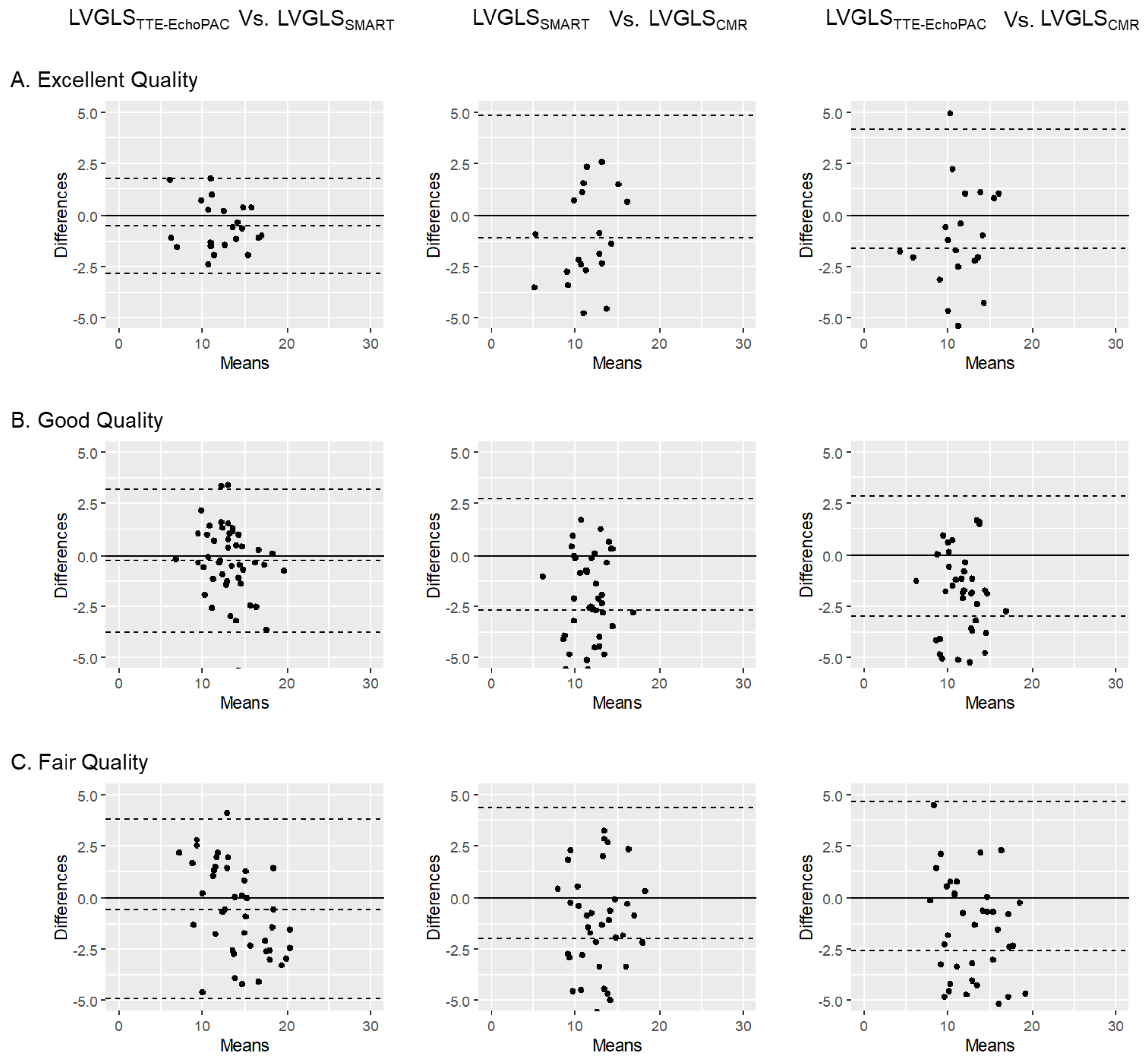

#### Supplemental Result 3. Subgroup analysis of LVM according to image quality

A. Excellent Quality

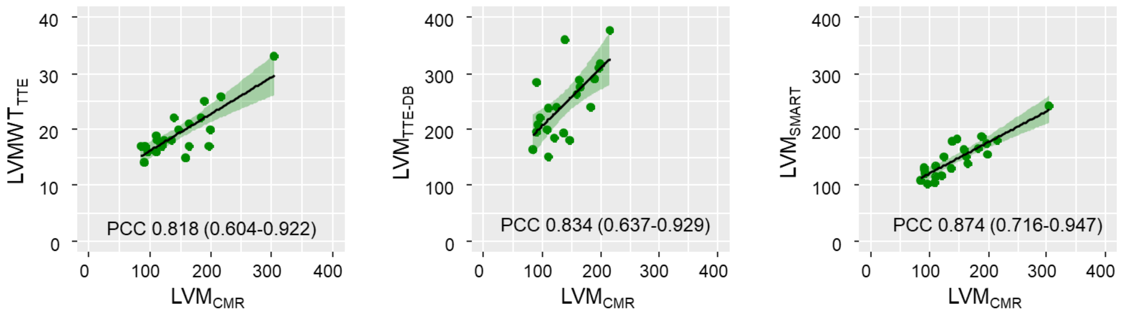


B. Good Quality

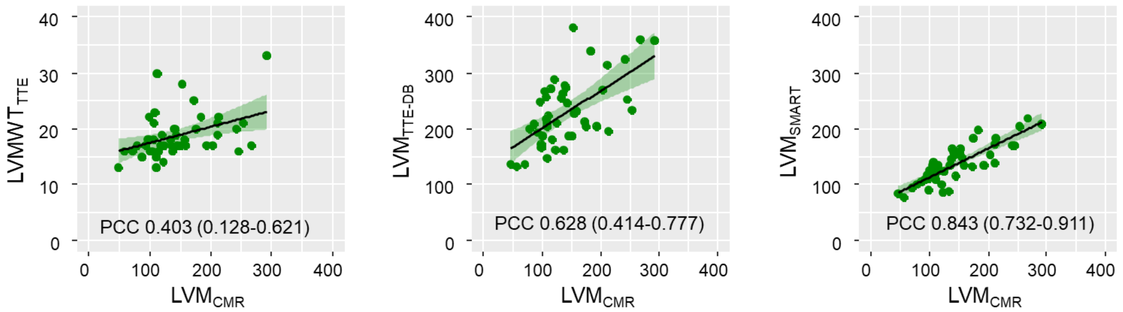


C. Fair Quality

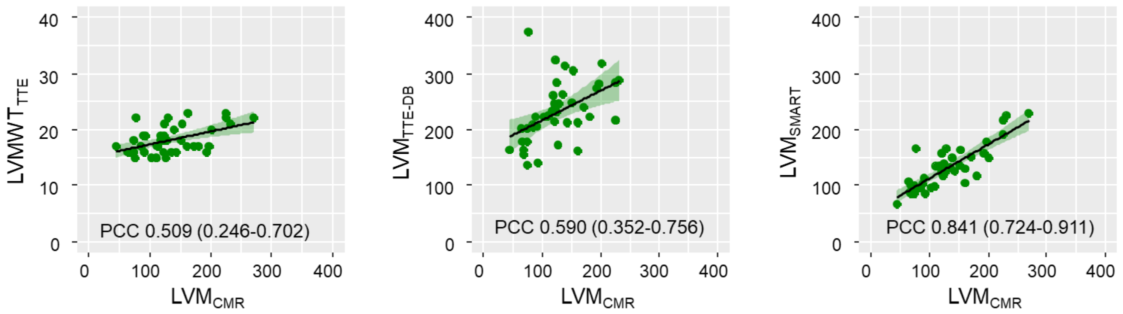

**Supplemental Result 4. Bland–Altman plots of LVM according to image quality**

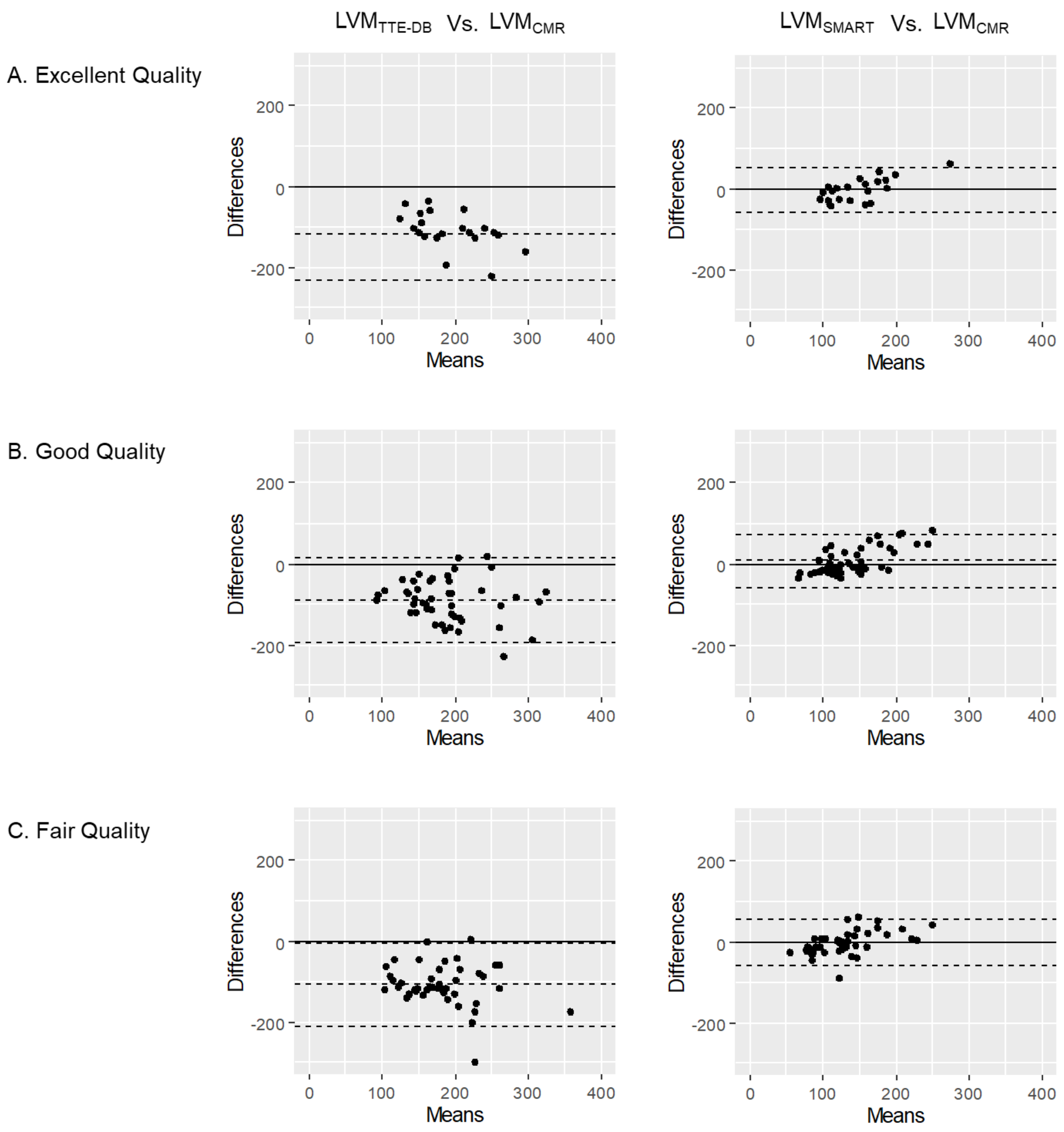

#### Supplemental Result 5. Subgroup analysis for LVGLS according to HCM phenotype

A. Septal

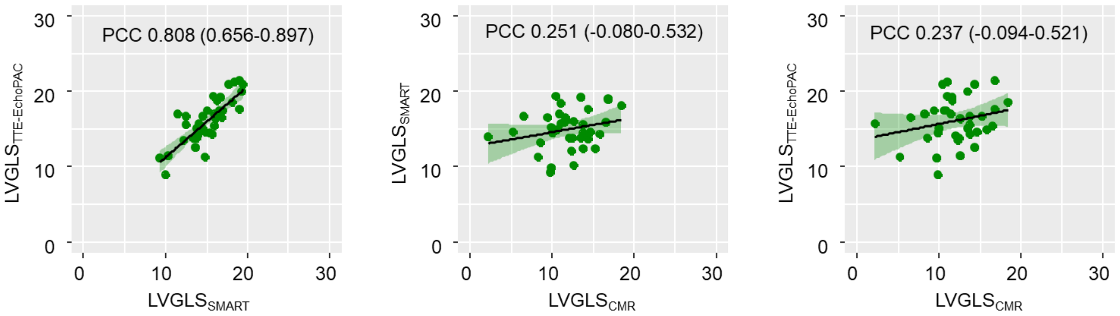


B. Mixed or Diffuse

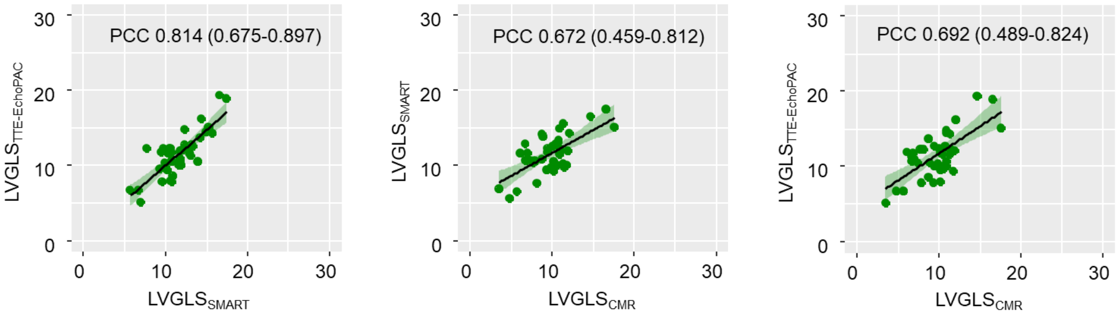


C. Apical

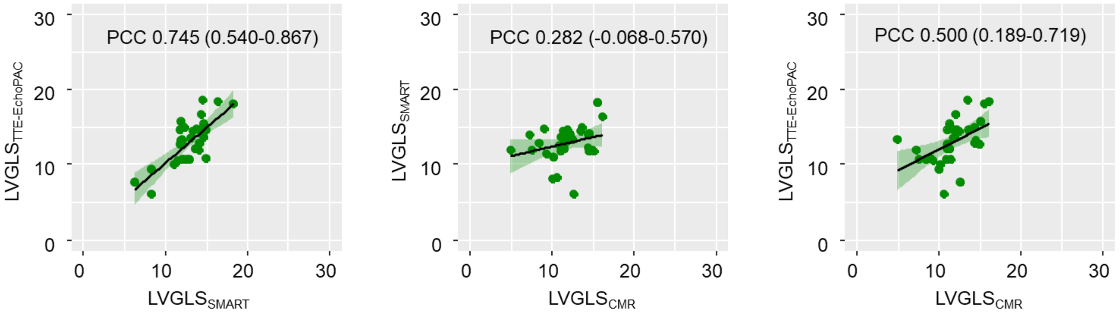

#### Supplemental Result 6. Subgroup analysis for LVGLS according to HCM phenotype

A. Septal

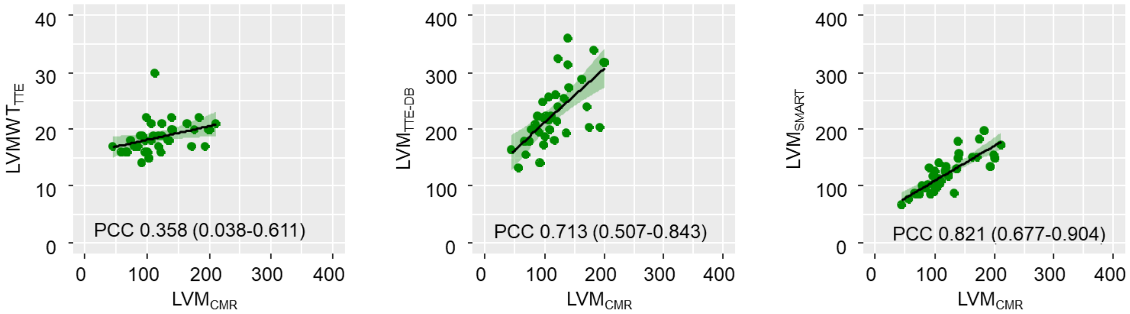


B. Mixed or Diffuse

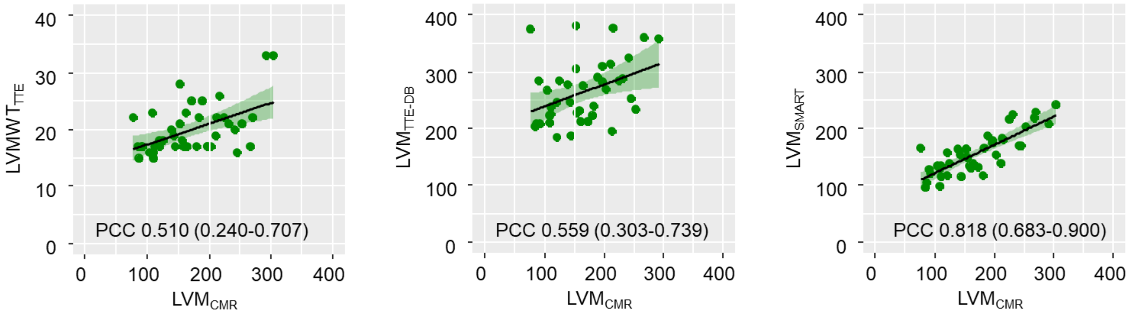


C. Apical

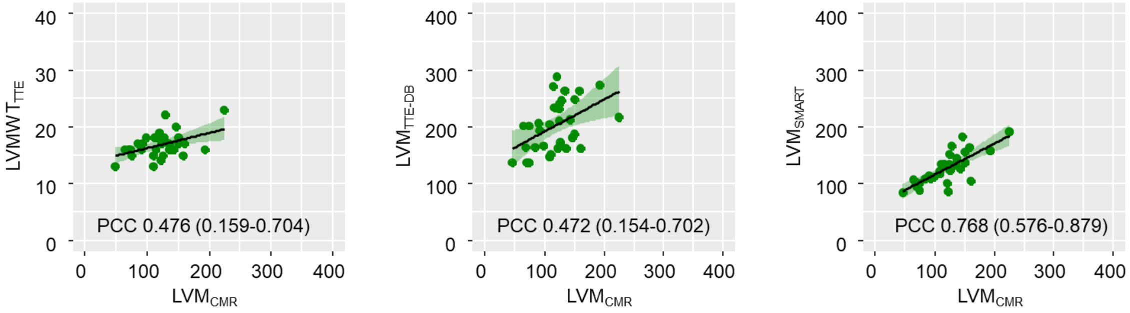

**References**

1. Jeon J, Ha S, Yoon YE, Kim J, Jeong H, Jeong D, et al. Echocardiographic view classification with integrated out-of-distribution detection for enhanced automatic echocardiographic analysis. arXiv [eess.SP; 2023]. Available from: https://arxiv.org/abs/2308.16483.

2. Jang Y, Choi H, Yoon YE, Jeon J, Kim H, Kim J, et al. An Artificial Intelligence-Based Automated Echocardiographic Analysis: Enhancing Efficiency and Prognostic Evaluation in Patients With Revascularized STEMI. Korean Circ J. 2024;54(11):743-56.

3. Starling MR, Walsh RA. Accuracy of biplane axial oblique and oblique cine angiographic left ventricular cast volume determinations using a modification of Simpson's rule algorithm. Am Heart J. 1985;110(6):1219-25.
